# High proportion of genome-wide homology and increased basal *pvcrt* levels in *Plasmodium vivax* late recurrences: a chloroquine therapeutic efficacy study

**DOI:** 10.1101/2021.01.22.21250265

**Authors:** Eduard Rovira-Vallbona, Nguyen Van Hong, Johanna H. Kattenberg, Ro Mah Huan, Nguyen Thi Huong Binh, Nguyen Thi Hong Ngọc, Pieter Guetens, Nguyen Luong Hieu, Nguyen Thị Thu Hien, Vu Thi Sang, Nguyen Duc Long, Erin Sauve, Tran Thanh Duong, Nguyen Xuan Xa, Annette Erhart, Anna Rosanas-Urgell

## Abstract

Chloroquine (CQ) is the first-line treatment for *Plasmodium vivax* malaria in most endemic countries. Monitoring *P*.*vivax* CQ resistance (CQR) is critical but remains challenged by the difficulty to distinguish real treatment failure from reinfection or liver relapse. Therapeutic efficacy of CQ against uncomplicated *P*.*vivax* malaria was evaluated in Gia Lai province, Vietnam. Sixty-seven patients were enrolled and followed-up for 42 days using microscopy and (RT)qPCR. Adequate clinical and parasitological response (ACPR) was 100% (66/66) on Day 28, but 75.4% (49/65) on Day 42. Eighteen recurrences (27.7%) were detected with a median time-to-recurrence of 42 days (IQR 35, 42) and blood CQ concentration <100ng/ml. Parasite genotyping by microsatellites, SNP-barcoding and whole-genome sequencing (WGS) identified a majority of homologous recurrences, with 80% (8/10) showing >98% identity-by-descent to paired Day 0 samples. Primary infections leading to recurrence occurred in younger individuals (median age for ACPR=25 years [IQR 20, 28]; recurrences=18 [16, 21]; p=0.002), had a longer parasite clearance time (PCT for ACPR*=*47.5h [IQR 36.2, 59.8]; recurrences=54.2h [48.4, 62.0]; p=0.035) and higher *pvcrt* gene expression (median relative expression ratio for ACPR*=*0.09 [IQR 0.05, 0.22]; recurrences=0.20 [0.15, 0.56]; p=0.002), but there was no difference in *ex vivo* CQ sensitivity. This study shows that CQ remained largely efficacious to treat *P*.*vivax* in Gia Lai, *i*.*e*. recurrences occurred late (>Day 28) and in the presence of low blood CQ concentrations. However, the combination of WGS and gene expression analysis (*pvcrt*) with clinical data (PCT) allowed to identify potential emergence of low-grade CQR that should be closely monitored.

## INTRODUCTION

*Plasmodium vivax* was responsible for 6.4 million malaria cases globally according to the World Health Organization (WHO) estimates for 2019, with most of them occurring in Asia (1). Historically mistaken as benign, *P. vivax* is becoming the predominant malaria species in countries where the incidence of *Plasmodium falciparum* is decreasing as they are moving towards malaria elimination (2). Whereas addressing asymptomatic infections and drug resistance are cross-cutting issues in the elimination of both *P. vivax* and *P. falciparum*, the treatment and control of *P. vivax* present additional unique challenges due to key biological differences, such as the presence of liver hypnozoites responsible for relapses (*i*.*e*. reactivation of dormant parasite forms leading to blood-stage infection), the low parasite density in peripheral blood, and the production of gametocytes (*i*.*e*. parasite transmissible stages) before the onset of symptoms (2,3).

Chloroquine (CQ) is the first-line drug treatment for *P. vivax* in most endemic countries (1). Chloroquine resistance (CQR) in malaria parasites emerged first in *P*.*falciparum* shortly after the start of widespread use as an antimalarial treatment in the 1950s (4). The first report of *P. vivax* CQR emerged 30 years later in Papua New Guinea and, since then, decreasing efficacy of CQ against *P. vivax* has been reported across malaria endemic areas around the world, although with different degrees of certainty (3,4). In countries where high level CQR has been confirmed (*i*.*e*. Papua New Guinea, Indonesia and Malaysia) artemisinin-based combination therapies have been adopted as the universal antimalarial first-line treatment instead of CQ (1,4).

In order to evaluate the efficacy of CQ regimens against *P. vivax*, the WHO recommends to conduct therapeutic efficacy studies (TES) with at least 28 days of patient follow-up after treatment administration (5). Recurrences within this period with a confirmed CQ blood concentration over the minimum effective concentration (MEC) of 100 ng/ml would support the presence of CQR. However, it has also been shown that a long-acting antimalarial drug like CQ can linger in the bloodstream for approximately 35 days and delay appearance of relapses up to six weeks (6,7). Extended follow-up periods of 42 or 63 days are therefore recommended to detect recurrences with parasites at early stage of CQR (5,8). Nonetheless, a main challenge of *P. vivax* TES studies is to distinguish a parasite recrudescence caused by real treatment failure from a relapse or new infection, for which additional *ex vivo* and *in vitro* laboratory tests are required (8). On one hand, genotyping of parasites at polymorphic genome regions can show whether recurrent parasite clones differ from those in the initial infection-especially when using high-resolution approaches at whole-genome level (9,10)-but if applied alone they remain limited to unambiguously differentiate a recrudescence from a genetically homologous relapse. On the other hand, short-term *ex vivo* cultures of isolates exposed to CQ and genotyping of drug resistance markers can be combined to characterize isolates CQR properties.

The identification and validation of CQR markers in *P. vivax* has unfortunately been hampered by the inability to long-term culture the parasite and evaluate adaptation to extended drug-exposure (11). Despite this limitation, numerous studies have linked CQR to both chloroquine resistance transporter (*pvcrt*) and multidrug resistance gene-1 (*pvmdr*1) genes. Associations with SNPs and/or copy number variation in both *pvcrt* and *pvmdr1* have remained weak and sometimes contradictory (12–16). At the transcriptional level, increased expression of *pvcrt* and *pvmdr1* was found in CQR infections from the Brazilian Amazon (17), whereas *pvcrt* expression was upregulated in a genetic cross of *P. vivax* parasites differing in their CQ response in experimental non-human primate infections (18).

In Vietnam, *P. vivax* accounts for 40% of all malaria cases, which occur mainly among ethnic minorities in forested areas in central and southern provinces (19). National guidelines for the treatment of *P. vivax* infection follows WHO recommendation of 3 days of CQ (25 mg/kg) followed by 14 days of primaquine (PQ; 0.25 mg/kg) radical-cure treatment to eliminate liver hypnozoites. To date, only one study conducted in Quang Nam province (Central Highlands) between 2009-2011 confirmed *P. vivax* CQR by reporting a 3.5% failure rate (n=8) at Day 28 and three cases with CQ blood concentrations >100ng/ml, although no genetic analysis of recurrences were performed (20). A few other reports d of suspected *P. vivax* CQR have been published in the last twenty years in Binh Thuan, Binh Phuoc and Ninh Thuan provinces (recurrence rates at Day 28 or later ranging 13%-57%), but CQR could not be confirmed due to the lack of either blood CQ levels or genotyping of recurrent infections (21)(22)(23).

In the present study we assessed the *in vivo* susceptibility of *P. vivax* to CQ in Gia Lai (Central Vietnam), the province with the highest malaria burden in the country in recent years (19,24). We used good manufacturing practice certified CQ and administered the currently recommended radical cure regimen of PQ at the end of the 42 days of follow-up. Importantly, we provide a thorough analysis of treatment failures by combining *ex vivo* drug sensitivity assays, three technical approaches to genetic analysis of recurrences, as well as genotyping of candidate markers of drug-resistance including gene expression analysis.

## MATERIALS AND METHODS

### Study site

The study was conducted between May 2015 and February 2017 in Chu R’Cam commune’s health center (CHC), Krong Pa district (Gia Lai province, Central Highlands, Vietnam), where a small field laboratory was set-up. In addition, CHC in the nearby communes of Ia R’Sai, Ia R’Suom and Uar were asked to refer all potentially eligible patients to the reference CHC. Chu R’Cam is located 120 km south-east of the provincial capital Pleiku, next to the Ba river in a large valley surrounded by hilly and partially forested areas (25). The local population is mainly from J’Rai ethnic minority, which has in general a low socio-economic status and education level. Local housing structures consist of government supported brick or wooden houses built on concrete floors. The main occupation is slash-and burn agriculture in forest fields, which often requires sleeping in the forest. The climate is tropical, with the dry season from November to April and the rainy season from May to October. Malaria transmission usually begins in May and peaks from September to December, with the lowest incidence between February and April. At the time of study initiation in 2015, Gia Lai province had reported 4367 microscopically confirmed malaria cases, the highest burden in the country, of which 1051 *P. falciparum* and 1124 *P. vivax* occurred in Krong Pa district (19,24). Malaria control by the provincial malaria station relies on early diagnosis by light microscopy (LM) or rapid diagnostic tests, annual distribution of long-lasting insecticidal treated nets, promotion of community-based prevention behavior and monitoring of drug resistance through regular TES. The presence of *P. falciparum* artemisinin resistance was confirmed in the province in 2017 (25).

### Study design and trial procedures

The study was designed as a 42-day drug efficacy study to evaluate clinical and parasitological responses after treatment of *P. vivax* uncomplicated malaria infections. Patients aged >1 year-old presenting to the CHC with fever (≥ 37.5° C) and/or history of fever during the previous 48 hours were screened for malaria infection by blood smears from finger prick, and hemoglobin (Hb) concentration was measured using Hb201+ System (Hemocue). Those with LM confirmed *P. vivax* mono-infection and asexual parasite density >250 parasites/µl were invited to enroll and provide written informed consent. Individuals diagnosed with *P. falciparum* or mixed infections, signs of severe malaria, febrile diseases other than malaria, or any chronic medical condition, as well as pregnant and breastfeeding women, were excluded. Prior to treatment, 5 ml of whole blood was collected by venepuncture for nucleic acid purification and *ex vivo* drug assays (200µl were transferred into an EDTA microtainer, 100 µl were transferred into an RNAprotect-containing containing tube and stored at −20°C, and remaining blood was processed as described below). After treatment initiation patients remained in the health center where they were monitored by LM every 12 hours for 72 hours (Day 3) or until parasite clearance was confirmed by two consecutive LM negative slides. On Days 7, 14, 28, 35 and 42 patients were asked to return for follow-up visits or were visited at home by health centre staff. At each visit, a finger prick was conducted to prepare thick and thin smears, 200μl of blood was transferred into an EDTA microtainer (for DNA extraction), and another 100µl was transferred into an RNAprotect-containing tube (for RNA extraction). On Day 7, 100μl of blood were collected onto chromatography paper for drug concentration measurements (31ETCHR, Whatman). Hb levels were measured on Days 14, 28, and 42. In case of confirmed recurrence, a 5ml venous blood sample and a blood spot on chromatography paper were collected prior to rescue treatment administration. A system of passive detection of fever cases was maintained throughout the study period.

### Treatment

All patients received a full treatment with CQ (Nivaquine®, Sanofi) at a total dose of 25mg of CQ base/kg body weight, given over 3 days and under direct observation (10mg the first two days and 5mg on the third)(26). Exact dosage given per patient was registered on case report forms. Individuals who vomited their medication within the first 30 minutes after treatment received another full dose, whereas those vomiting between 30 and 60 minutes after treatment received another half dose. The time when patients received the first dose was defined as Day 0. PQ (0.25mg/kg/day for 14 days, Sanofi/Valeant Pharmaceuticals) was given at the end of follow-up on Day 42 to all patients testing negative for glucose-6-phosphate dehydrogenase deficiency (G6PD; CareStart, Access Bio) under supervision of health workers. Patients with recurrences at Day 42 or earlier were administered radical cure treatment as per national guidelines, consisting of CQ (25mg/kg, 3 days) together with PQ (0.25mg/kg/day for 14 days)(26).

### Microscopy

Malaria diagnosis by LM was conducted by examining thick and thin smears stained with freshly prepared 10% Giemsa for 15 min under 1000X magnification. Parasite density was calculated by counting the number of asexual parasites per 200 white blood cells (WBC) with a hand tally counter and expressed as parasites/μl of blood assuming a WBC density of 8000 cells per μl. Blood smears were considered negative when no asexual parasites were found after counting 1000 WBC. All smears were read in duplicate by two trained microscopists and the final density was expressed as the average result. A third reading was conducted by another technician in case of discrepancy in positivity, species diagnosis, or parasite count by >25%. Fifteen per cent randomly selected blood smears were sent to ITM (Antwerp, Belgium) for external quality control.

### *P. vivax* quantification by (RT)qPCR

DNA was extracted from 200 µl whole blood in EDTA using Favorprep 96-well Genomic DNA kit (Favorgen) and eluted in 200 µl of water. Duplex qPCR targeting *P. vivax* and *P. falciparum* 18S ribosomal RNA genes was performed in samples from Day 0 using TaqMan Universal Master Mix II and StepOne Plus Real-Time PCR System (Applied Biosystems) (27). The same method was adapted as a *P. vivax* monoplex qPCR and used for *P. vivax* quantification in follow-up samples. RNA was extracted from RNAprotect samples collected at Day 0 and from all follow-up samples positive in the *pv18S rRNA* qPCR, using RNeasy Plus 96-kit (Qiagen) (25). Final RNA elution was conducted in 100 µl of RNAse-free water, and the presence of quality RNA was confirmed in a random selection of 80 samples by one-step reverse transcription qPCR (RTqPCR) targeting *pv18S rRNA* transcripts, using LightCycler Multiplex RNA Virus Master mix in a LightCycler480 thermocycler (Roche). The presence of *P. vivax* stage V gametocytes was determined by RTqPCR amplification of *pvs25* RNAs (28). A standard curve (10^6^-1 copies/µl) of plasmids containing the *pvs25* PCR fragment was included in each plate for copy number quantification (28).

### *Ex vivo* schizont maturation assays (SMA)

Venous blood vacutainers were centrifuged, and plasma and buffy coat stored at −20°C. Infected red blood cells (iRBC) were diluted to 25% haematocrit in sterile PBS and passed through an autoclaved 10ml syringe (Terumo, SS10LE1) prefilled with cellulose powder 2.5ml-layer (Sigma-Aldrich, C6288) on top of a lens-cleaning paper to remove white blood cells. In all samples with a ring-stage proportion ≥65%, 150 µl iRBC were separated for *ex vivo* drug sensitivity SMA and remaining iRBC pellet was cryopreserved in liquid nitrogen or stored at −20°C. SMA was performed by adapting WHO microtest, as previously described (25,29). A stock solution of 640µM CQ in water (Sigma-Aldrich, C6628) was pre-dosed in seven 2-fold serial dilutions in water (800-12.5nM), and 25 µl/well were distributed in a 96-well plate, air-dried overnight inside a laminar-flow cabinet and stored at 4°C (30). The 150µl iRBC were resuspended to 2% hematocrit in pre-warmed McCoy’s 5A medium (Gibco) supplemented with 20% human serum (non-exposed Vietnamese individuals), and 200 µl/well of the iRBC-medium mixture were added to pre-dosed plates, including drug-free wells. Plates were cultured at 37°C and drug-free wells monitored by LM at 34h, and thereafter every 2-4h until the number of schizonts reached 40% (or until 42 hours). The percentage of schizonts per each drug concentration was determined by double-reading of blood smears and result expressed as schizonts % relative to the drug-free well. Half maximal inhibitory concentrations (IC50) were calculated using WWARN’s online In Vitro Analysis and Reporting Tool (31).

### Drug concentration in blood

Concentrations of CQ and desethyl-chloroquine (DCQ, *i*.*e*. the main chloroquine metabolite) were measured in paired samples on Day 7 and Day of recurrence (DRec). Blood expelled onto chromatography papers was air dried and stored in separate plastic bags with silica gel. Drug measurements were conducted by liquid chromatography–mass spectrometry at the Department of Clinical Pharmacology, Mahidol-Oxford Tropical Medicine Research Unit (Bangkok, Thailand). The minimum effective concentration (MEC) was considered a [CQ]+[DCQ] value of 100 ng/ml (8).

### Microsatellites genotyping

Paired Day 0 and DRec samples were genotyped at polymorphic markers Pvmsp1F3, MS4, MS10 and PvSal1814, selected based on the high degree of heterozygosity found in Central Vietnam and adapting previously published protocols (see Table S1) (32) (33,34). All products were run on a 2% agarose gel, and 20μl pool of Pvmsp1F3 (5μl), MS4 (7.5μl) and MS10 (7.5 μl) PCR products and PvSal1814 products were sent off for capillary electrophoresis at Genoscreen (Lille, France). Allele calling was performed using GeneTools (Syngene) and GeneMarker v2.4.0 (Softgenetics). All samples were double-checked manually and the predominant allele (*i*.*e*. the highest peak) and minor alleles within at least two-thirds of the height of the predominant allele were scored. Alleles were considered identical if size difference was ≤2 bp. Complexity of infection (COI; *i*.*e*. the estimated number of genetically distinct clones within an infection) was defined as the highest number of alleles found in any of the four markers. Genotypes at Day 0 and DRec were classified as homologous (when all or some of the alleles present in Day 0 sample were found in DRec sample for all markers), heterologous (when none of the alleles present in Day 0 sample were found at DRec for at least one marker), or indeterminate (when no amplification was achieved for any marker).

### SNP barcode

A molecular barcode consisting of 38 SNPs across the *P. vivax* genome was used to genotype samples in the context of MalariaGEN SpotMalaria Project (Welcome Sanger Institute, Hinxton, UK), using MALDI-TOF mass spectrometry of PCR amplicons in a MassARRAY® System (Agena BioScience) (25)(35). Nucleotide sequence results were provided in the form of a Genetic Report Card (see Data S1). Recurrences were classified as homologous to Day 0 if there were no nucleotide differences in any non-heterozygous position of the barcode, irrespective of the total number of successfully genotyped SNPs.

### Whole-genome sequencing

DNA was initially subject to selective whole-genome amplification (sWGA) with *phi29* DNA polymerase (New England Biolabs) according to previously published methodology (30 µl of DNA in 50 µl reaction volume)(36). Product was purified with AMPure XP beads (Beckman-Coulter) and 15µl were used in a second sWGA round (50 µl reaction volume). Final sWGA purified product concentration was determined in a Qubit 2.0 Fluorometer (Life Technologies), and a minimum of 1mg was sent to BGI (Hong Kong) for library preparation and 150bp paired-end WGS on a HiSeq X Ten (Illumina). FASTQ files were trimmed using Trimmomatic to remove adapters and low quality reads, and aligned to the reference genome *PvP01* using the BWA-MEM algorithm v0.7.17 (37). Variants were called using HaplotypeCaller in GVCF mode followed by Joint-Call Cohort (GATK 4.1.2.0, Broad Institute) and filtered to include only biallelic SNPs in the core genome, MQ>50, QUAL>30 and combined DP ≥100, resulting in 191849 high quality SNPs (38). Samples with at least 10X coverage were kept for subsequent analysis (see Table S2). Within-host infection complexity was assessed using within-sample F statistic (FWS) and FWS>0.95 was considered as a proxy for a monoclonal infection (39,40). Pairwise comparisons between all samples were analysed in as identity-by-state (IBS) and identity-by-descent (IBD). For the IBS approach, the proportion of discordant SNPs was determined by calculating the Prevosti distance (*i*.*e*. number of allelic differences/number of possible differences). Distance was fitted as the sum of four Gaussian distributions and the mean of the lowest distribution was set as the threshold for IBS-homologous recurrence (see Fig S1). For the IBD analysis, PED and MAP file formats were created from using VCFtools and the proportion of the genome IBD between pairs of samples was calculated using isoRelate R package (41). Genetic distance was calculated using estimated mean map unit size from *P. chabaudi chabaudi* of 13.7 kb/cM (42). We set thresholds of IBD on the minimum number of SNPs (20) and length of IBD segments (50.000) reported to reduce false positive calls. Pairwise comparisons with ≥95% of their genome identical-by-descent were considered homologous and all pairwise comparisons with shared IBD <95% were considered heterologous.

### Molecular markers of drug resistance

Gene expression of *pvcrt* and *pvmdr1* were determined directly on RNA samples by one-step RTqPCR. *β-tubulin* was used as reference gene. Each 10µl reaction volume contained 2µl of LightCycler EvoScript RNA SYBR Green I Master (Roche), 0.4µM of forward and reverse primers (from ref. (17)), and 2.5µl of RNA template. LightCycler480 (Roche) thermal cycling conditions were 60°C for 15 minutes, 95°C for 10 minutes, 45 cycles of 95°C for 10 seconds and 58°C for 30 seconds, followed by a melting curve step to ensure amplification specificity. Samples were run in triplicate and excluded from calculations if final SD of duplicates after removal of outliers was >0.4. Gene expression was estimated using the efficiency-adapted Pfaffl relative quantification model (43), in which the relative expression ratio (R) is calculated as the efficiency of the target genes (see Fig S2) raised to the power of the Ct difference between a given sample and a control (*i*.*e*. mean Ct + 2*SD of all Day 0 samples with ACPR), and divided by the same formula applied to reference gene *β-tubulin*. Mutations in *pvmdr1* codons 976 (Y→F) and 1076 (F→L) were genotyped by nested PCR amplification using primers from *Golassa et al* (primary) and *Lin et al* (nested; see Table S1) (44,45). The 646bp amplification products were sequenced at Genoscreen (Lille, France). Nucleotide sequences (Genbank accession numbers MW245736-MW245808) were entered to BioEdit 7.0 and aligned using ClustalW with default parameters.

### Clinical trial endpoints

The primary endpoint was adequate clinical and parasitological response (ACPR) at 28 and 42 days of follow-up. Treatment failures were classified as early treatment failure, late clinical failure (LCF, *i*.*e*. parasitemia with fever or signs of severe malaria between Day 4 and Day 42) or late parasitological failure (LPF, *i*.*e*. parasitemia between Day 7 and Day 42), based on WHO guidelines (5). Patients who discontinued due to reappearance of parasites and subsequent rescue treatment were considered evaluable for Day 42 endpoint analysis. Secondary endpoints were parasite clearance estimates obtained from the log_e_ parasitemia-time LM data entered into WWARN Parasite Clearance Estimator (46), asexual and sexual parasite carriage on Day 0-3 and during follow-up (including submicroscopic) and severe anemia (Hb<7g/dl) on Day 7.

### Statistical analysis

For sample size calculation, the initial estimation of 5% treatment failure rate (based on the Quang Nam study (20)) was amended to 20% upon completion of the first year of the study, after observing treatment outcome results together and an extremely low malaria incidence in the district. To detect a minimum failure rate of 20%, a minimum sample size of 61 patients was necessary (10% precision and 95% confidence level), which was extended to 67 to allow for 10% loss to follow-up. All clinical data were double entered into a Microsoft Access database and transferred to Stata v11.0. The primary study outcomes were analysed using both *per-protocol* and intention-to-treat (ITT) patient population. Comparisons between groups for continuous variables were performed using Wilcoxon rank-sum test and proportions were compared using Chi-square or Fisher’s exact tests, when appropriate. Correlation between variables was assessed by Spearman’s test. Clearance time for total parasites and for gametocytes was estimated using Kaplan–Meier survival curves and comparisons between subgroups were done using log-rank test. Time was computed in hours and since treatment administration. Gene expression data was log-transformed, and comparisons between groups were performed by pairwise Wilcoxon rank-sum test, with Benjamini & Hochberg correction in cases of multiple testing. A linear model adjusted for the proportion of ring-stage parasites was applied to investigate the potential effect of stage composition on gene expression (47). Paired analyses between Day 0 and DRec were done using Wilcoxon matched-pairs signed-rank test. Statistical analysis and graphs were performed in Stata v11.0, R v4.0 or GraphPad Prism (v9.0). *P*-values <0.05 were considered statistically significant.

### Ethics approval and consent to participate

The study received approval from ethics committee of the NIMPE-Ministry of Health, Hanoi, Vietnam (351/QD-VSR and QD2211/QD-BYT); the Institutional Review Board of the Institute of Tropical Medicine, Antwerp, Belgium (937/14); and the ethics committee of Antwerp University Hospital (UZA), Antwerp, Belgium (14/15/183). Written informed consent was obtained from all participants or their respective parents/guardians in case of minors. The trial was registered on ClinicalTrials.gov under identifier NCT02610686.

## RESULTS

### Patient recruitment and characteristics at baseline

A total of 480 patients were screened at the different CHC out of which 68 (14.2%) had *P. vivax* infection confirmed by LM. Sixty-seven were enrolled and completed CQ treatment (Fig 1). Twenty-seven (40.3%) patients self-presented at Chu R’Cam CHC, whereas the other 40 (59.7%) were referred from nearby communes. The age range was 8 to 45 years old and most patients were male (91%; Table 1). Two thirds of participants reported having had an episode of malaria in the past year (62.7%).

**Table 1.** Patients characteristics at baseline.

| Variable | N=67 |
| --- | --- |
| Age group: |  |
| years, median (IQR) | 22 (18, 26) |
| >1-15 years, n (%) | 6 (9.0) |
| >15-25 years, n (%) | 37 (55.2) |
| >25 years, n (%) | 24 (35.8) |
| Sex, males (%) | 61 (91.0) |
| Ethnic group, n (%) |  |
| J'Rai | 52 (77.6) |
| Kinh | 15 (22.4) |
| Weight (kg), median (IQR) | 52.5 (48, 61) |
| Had malaria in the previous year, n (%) | 42 (62.7) |
| Took antimalarials in the previous 14 days | 0 (0) |
| G6PD deficiency | 1/67 (1.5) |
| Fever at enrolment ($\geq 37.5^{\circ}\text{C}$), n (%) | 56 (83.6) |
| Body temperature: |  |
| All ($^{\circ}\text{C}$), median (IQR) | 38.5 (37.5, 39.5) |
| Febrile only ($^{\circ}\text{C}$), median (IQR) | 39.0 (38.4, 39.5) |
| Clinical symptoms: |  |
| Headache, n (%) | 67 (100.0) |
| Fatigue, n (%) | 66 (98.5) |
| Dizziness, n (%) | 48 (71.6) |
| Chills, n (%) | 37 (55.2) |
| Sweats, n (%) | 21 (31.3) |
| Nausea, n (%) | 17 (25.4) |
| Pain, n (%) | 6 (9.0) |
| Vomiting, n (%) | 6 (9.0) |
| Diarrhea, n (%) | 3 (4.5) |
| Hemoglobin (g/dL), median (IQR) | 13.5 (12.5, 14.8) |
| Anemia (Hb <11g/dL), n (%) | 7 (10.4) |
| Parasitological data by LM: |  |
| Asexual parasites/$\mu\text{l}$, geomean (95% CI) | 5992 (4594, 7815) |
| Ring-stage %, median (IQR)* | 75 (43, 91) |
| Gametocyte prevalence, n (%) | 58 (86.6) |
| Gametocytes/$\mu\text{l}$, geomean (95% CI) | 223 (171, 290) |

\*N=64.
Hb, hemoglobin; G6PD, glucose-6-phosphate dehydrogenase.

**Fig 1.**
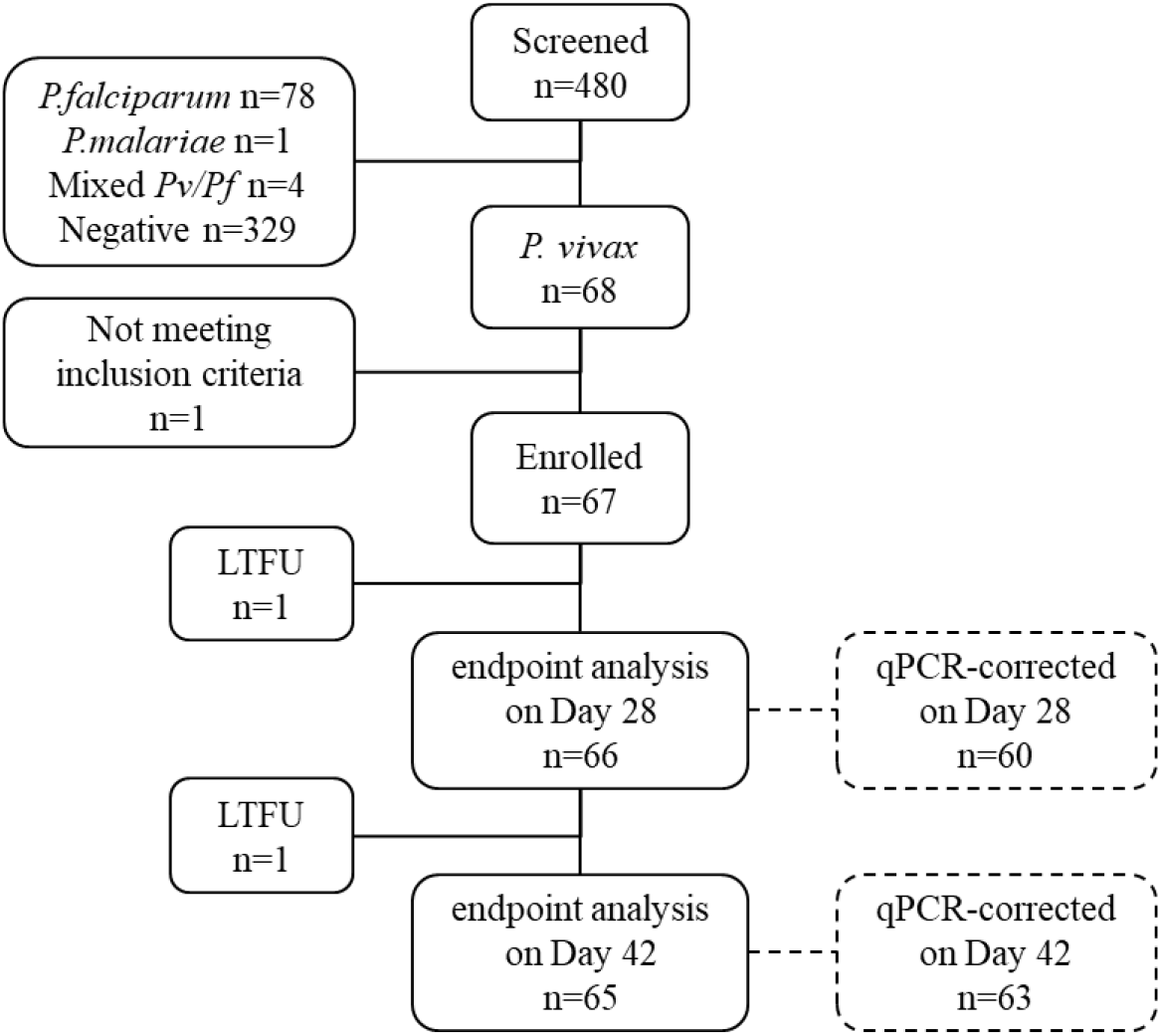
Flow chart of patient screening, enrolment, and follow-up. *LTFU*, loss to follow-up.

Fever was detected in 56 patients (83.6%) at the time of enrolment, 7 patients (10.4%) presented anemia (Hb<11g/dL) and 1 patient was detected with G6PD deficiency. All patients reported headache and fatigue together with other clinical symptoms, most commonly dizziness and chills. Up to 87% of infections carried detectable level of gametocytes by LM (Table 1). Asexual parasite density was significantly correlated with body temperature (*ρ*=0.435, p<0.001), density of gametocytes (n=58; *ρ*=0.525, p<0.001) and proportion of ring stages (n=64; *ρ*=0.431, p<0.001) but did not differ between age groups (p=0.971).

### CQ efficacy endpoints

Overall, CQ treatment was well tolerated, with 3 patients reporting moderate adverse effects (dizziness, diarrhea, and abdominal pain). The median CQ dose received by patients was 24.5 mg/kg (IQR 24.0-25.4). Endpoint analysis was conducted using LM data. PCT and parasite clearance half-life (PC_1/2_) were estimated at 48.3 h (maximum 95.3 h) and 4.1 h (max.13 h), respectively (Table 2). All patients were negative by LM at Day 4. Sixty-six (98.5%) and 65 (97.0%) patients completed the 28-day and the 42-day follow-up, respectively (Table 2). At Day 28, the rate of ACPR was 100% (98.5% [66/67] based on ITT analysis), whereas at Day 42 the rate of ACPR was 75.4% (73.1% [49/67] based on ITT analysis). Out of 16 recurrences, three presented with fever (LCF, days 33-42), while the other 13 were afebrile (LPF, >Day 35). Mean parasitemia at recurrence was 538.7 parasites/μl (95% CI 227.8, 1274.0), and was higher in LCF (n=3; 3024.2 parasites/μl [95% CI 1946.9, 4697.6]) compared to LPF (n=13; 361.7 [95% CI 143.5, 911.9]; p=0.026). Gametocytes were observed in 6/16 (37.5%) infections at DRec.

**Table 2.** Study endpoints.

| Endpoints | n/N (%) or median (IQR) |
| --- | --- |
| **Primary:** |  |
| ACPR |  |
| Day 28 | 66/66 (100.0) |
| Day 42 | 49/65 (75.4) |
| Treatment failures (Day 42): |  |
| LCF | 3/65 (4.6) |
| LPF | 13/65 (20.0) |
| Time to recurrence in days | 42 (35, 42) |
| **Secondary:** |  |
| PCT, hours (n=66) | 48.3 (36.8, 60.1) |
| PC<sub>1/2</sub>, hours (n=63) | 4.1 (3.5, 5.4) |
| Patients with asexual parasitemia at: |  |
| Day 1 | 56/67 (83.6) |
| Day 2 | 25/67 (37.3) |
| Day 3 | 2/66 (3.0) |
| Patients with gametocytes at: |  |
| Day 1 | 29/67 (43.3) |
| Day 2 | 8/67 (11.9) |
| Day 3 | 0/66 (0) |
| Severe anemia (Hb<7g/dL; day 7) | 0/34 (0) |

ACPR, Adequate clinical and parasitological response; LCF, late clinical failure; LPF, late parasitological failure; PCT, parasite clearance time; PC<sub>1/2</sub>, parasite clearance half-life, *i.e.* time to decrease initial parasitemia by half in log-linear phase; Hb, hemoglobin.

After qPCR correction, parasite clearance rate did not differ significantly from that determined by LM (p=0.338, log-rank; Fig 2A), although the number of Day 3 positives increased to 13.9% (9/65) by qPCR as compared to 3% (2/65) by LM. On the other hand, gametocyte clearance rate was significantly slower by RTqPCR than by LM (p<0.001, log-rank; Fig 2B), and 11.3% were found gametocyte-positive on Day 3. Two additional recurrences were detected by molecular methods on Day 42 (total recurrences=18), resulting in a qPCR-corrected ACPR of 73% at the end of the 42-day follow-up (68.7% [46/67] as per ITT analysis).

**Fig 2.**
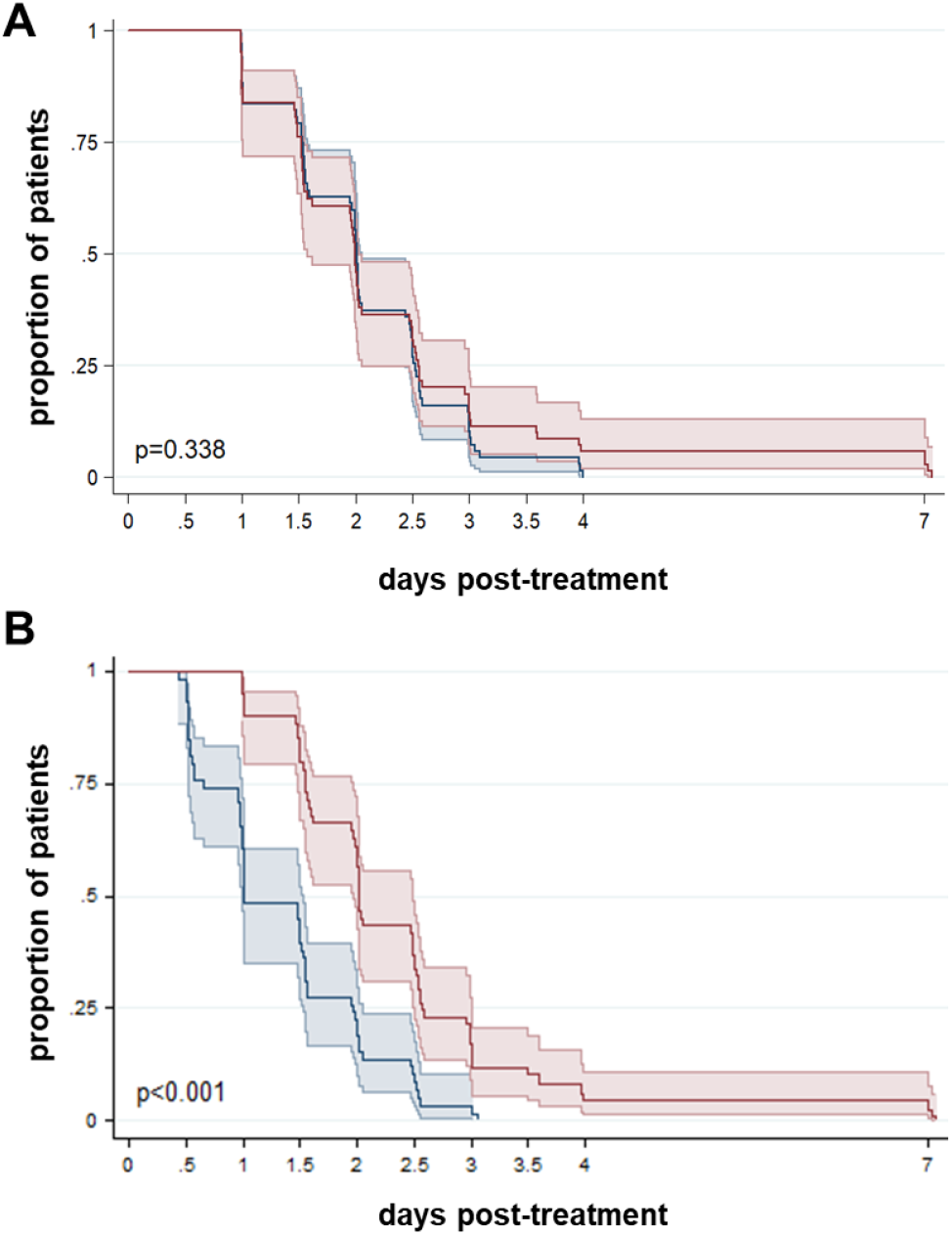
Time to parasite clearance. Kaplan-Meier survival curves for total parasite clearance (A) and gametocyte clearance (B). Results are stratified by light microscopy (n=67, blue line) and qPCR (A; n=63, red line) or RTqPCR (B; n=59, red line), respectively. Colored areas indicate 95% confidence intervals. P-values were calculated using log-rank test. One patient with no Day 7 sample by qPCR/RTqPCR was excluded from this analysis.

All 18 recurrences occurred after day 33 (median 42 days [IQR 35-42]; mean 38.7; range 33-43 days) and affected younger individuals (median age 18 years old [IQR 16, 21]), as compared to patients with ACPR (25 years [IQR 20, 28], p=0.002; Table 4). Recurrent and non-recurrent infections did not differ in the CQ dose administered (24.6 mg/kg [24.0, 25.5] *versus* 24.5 mg/kg [IQR 24.0, 25.0]; p=0.787). Blood concentrations of CQ and DCQ at DRec were found to be below 100ng/ml (median 30.3 ng/ml [IQR 15.2, 36.4]; max. value 52.3ng/ml) in the 14 patients with valid test result (all Day 7 concentrations were >100ng/ml: 502 ng/ml [IQR 358, 601]; n=15). PCT was longer in infections that presented recurrence (n=18; 54.2h [IQR 48.4, 62.0]) compared to those with ACPR (n=46; 47.5h [IQR 36.2, 59.8], p=0.035; Table 4), and a similar trend was observed for PC_1/2_ (recurrent: 4.8h [IQR 4.0, 5.5]; ACPR: 4.0h [IQR 3.4, 5.2], p=0.078). Asexual parasitemia at Day 0 was not significantly different between recurrent (n=49; mean 6762.3 p/ul [95% CI 3811.7, 11997.0]) and non-recurrent infections (n=16; 7114.3 p/ul [95% CI 3726.7, 13581.3], p=0.462), and neither was positivity at Day 2 (p=0.409) or Day 3 (p=0.772; Table 4).

### Genotyping of recurrent infections

Treatment failures were corrected by genotyping approaches and classified as genetically homologous or genetically heterologous infections relative to the initial infection at Day 0, using three technical approaches in increasing order of resolution. Initially, Day 0/DRec sample pairs were genotyped at four MS markers by PCR and capillary electrophoresis (Table 3 and Table S3). Overall, 2/18 (11%) samples at Day 0 and 4/15 (27%) at DRec were monoclonal. Fifteen sample pairs had data for at least 2 markers (Table 3). Median COI was 3 (IQR 2-3) at Day 0 compared to 2 (IQR 1-3) at DRec (p=0.080, signed-rank). Homologous recurrences by MS accounted for 80% (12/15) of total recurrences.

**Table 3.** Genotyping and categorization of recurrent infections. An extended version is provided in Supplemental Materials as Table

| Patient code | Microsatellites |  | SNP barcode |  | Whole-genome sequencing |  |  |  |
| --- | --- | --- | --- | --- | --- | --- | --- | --- |
|  | Discordant MS | Type | Discordant SNP | Type | IBS |  | IBD |  |
|  |  |  |  |  | Discordant SNP | Type | % IBD | Type |
| 003 | 0/3 | **homologous** | 1/38 | heterologous | - | - | - | - |
| 007 | 0/4 | **homologous** | 2/38 | heterologous | 14.3% | heterologous | 35.2% | heterologous |
| 008 | 0/4 | **homologous** | 0/37 | **homologous** | 2.2% | **homologous** | 99.9% | **homologous** |
| 010 | - | - | 1/38 | heterologous | - | - | - | - |
| 018 | 0/2 | **homologous** | - | - | - | - | - | - |
| 021 | 4/4 | heterologous | - | - | - | - | - | - |
| 022 | 0/4 | **homologous** | 0/37 | **homologous** | 6.8% | heterologous | 98.1% | **homologous** |
| 027 | 0/4 | **homologous** | - | - | 2.3% | **homologous** | 99.5% | **homologous** |
| 028 | - | - | - | - | - | - | - | - |
| 034 | ¼ | heterologous | 0/38 | **homologous** | 1.6% | **homologous** | 99.8% | **homologous** |
| 037 | - | - | - | - | 2.0% | **homologous** | 99.7% | **homologous** |
| 043 | 0/3 | **homologous** | 1/38 | heterologous | 12.6% | heterologous | 99.8% | **homologous** |
| 044 | 0/3 | **homologous** | - | - | - | - | - | - |
| 047 | 1/3 | heterologous | 0/22 | **homologous** | 8.0% | heterologous | 32.5% | heterologous |
| 048 | 0/3 | **homologous** | 0/20 | **homologous** | 1.7% | **homologous** | 99.5% | **homologous** |
| 050 | 0/3 | **homologous** | - | - | - | - | - | - |
| 053 | 0/4 | **homologous** | 0/38 | **homologous** | - | - | - | - |
| 054 | 0/4 | **homologous** | 5/38 | heterologous | 2.4% | **homologous** | 100% | **homologous** |
| **Homologous rate:** | 12/15 (80%) |  | 6/11 (55%) |  | 6/10 (60%) |  | 8/10 (80%) |  |

MS, microsatellites (PvmSP1F3+MS4+MS10+PvSal1814); IBS, identity-by-state; IBD, identity-by-descent.

In a second step, recurrences were genotyped using a 38-SNP molecular barcode (35). Barcodes were successful in 11 Day 0/DRec sample pairs, although two of them at only 20 and 22 SNP positions, respectively (Table 3 and Data S1). COI was 1 (IQR 1-1) at Day 0 and 2 (IQR 1-2) at DRec (p=0.034, signed-rank). Homologous recurrence rate was 55% (6/11). Compared to MS (n=10 pairs), the SNP-barcode identified 4 additional heterologous infections and categorized two MS-heterologous infections as homologous.

Finally, samples were subject to WGS aiming for a maximum level of resolution. Fourteen Day 0/DRec DNA sample pairs generated sequencing reads in both samples. Mean sequencing coverage ranged from 1.6–50.1 (overall mean 30.3 ± 14.5) and there was no significant difference in coverage between samples from Day 0 and DRec (p=0.280, paired t-test; Table S2). COI was 1 in 58% (7/12) of samples at Day 0 and 73% (8/11) at DRec. WGS analysis of identity-by-state (IBS; *i*.*e*. number of discordant SNP) identified 60% (6/10) of homologous recurrences (Table 3). Compared to MS (n=9 pairs), IBS categorized three originally homologous recurrences as heterologous. WGS data analysis of identity-by-descent (IBD; *i*.*e*. considering inheritance from a common ancestor) identified 80% (8/10) recurrent infections as homologous, all of them with >98% of their genome identical-by-descent with the paired Day 0 sample (median 99.7% [IQR 0.3%], n=8). Median IBD found between Day 0/DRec pairs (99.6% [IQR 98.4-99.8%], n=10) was significantly higher than the IBD found in all between-patient comparisons (0% [IQR 0-0.60%], n=196, p<0.001, Mann-Whitney U Test), suggesting it is unlikely that high IBD observed is caused by independent infections with a closely related clone. Results between IBS and IBD were discordant in 2 out of the 10 sample pairs (patients 022 and 043), which had a considerably high number of discordant SNP by IBS (6.8% and 12.6%) but high IBD proportion (98.1% and 99.8%); mean sequencing coverage was >34.2 for both samples and timepoints. Assuming IBD provides the highest level of resolution and most restricted definition of homology, the final rate of homologous recurrence (IBD-homologous) in the study was of at least 12% (8/65) out of all patients that completed the 42-day of follow-up, and of at least 44% (8/18) out of total recurrences.

Compared to patients with ACPR, patients with IBD-homologous recurrences (n=8) were not different in CQ uptake (24.1 mg/kg [IQR 23.0, 25.2], p=0.565) or parasite density at Day 0 (p=0.142; Table 4). However, median PCT among IBD-homologous was significantly longer than in those with ACPR (p=0.019).

**Table 4.**
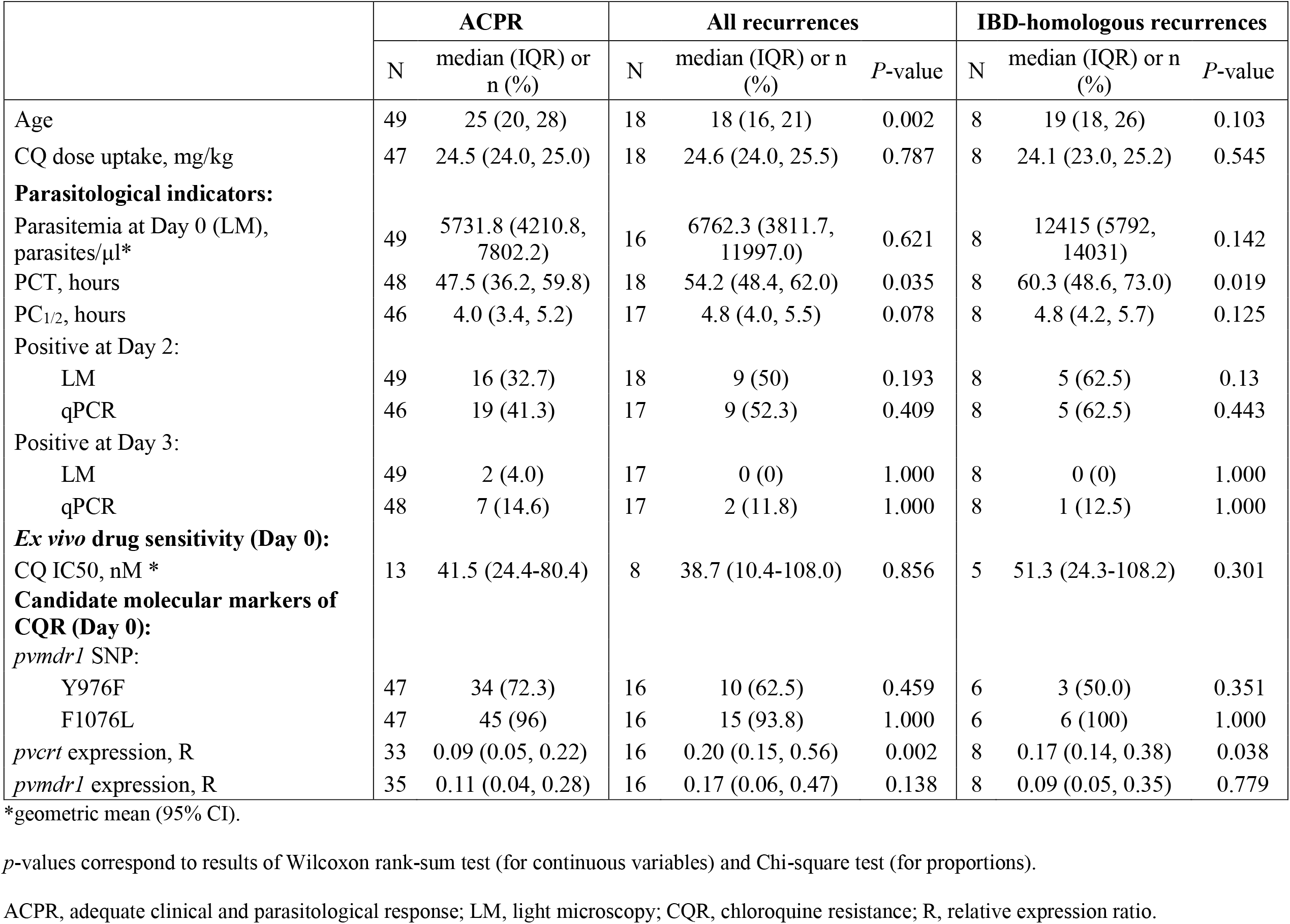
Main parasitological and drug-resistance characteristics of *P. vivax* infections, by treatment outcome at baseline.

|  | ACPR |  | All recurrences |  |  | IBD-homologous recurrences |  |  |
| --- | --- | --- | --- | --- | --- | --- | --- | --- |
|  | N | median (IQR) or n (%) | N | median (IQR) or n (%) | P-value | N | median (IQR) or n (%) | P-value |
| Age | 49 | 25 (20, 28) | 18 | 18 (16, 21) | 0.002 | 8 | 19 (18, 26) | 0.103 |
| CQ dose uptake, mg/kg | 47 | 24.5 (24.0, 25.0) | 18 | 24.6 (24.0, 25.5) | 0.787 | 8 | 24.1 (23.0, 25.2) | 0.545 |
| **Parasitological indicators:** |  |  |  |  |  |  |  |  |
| Parasitemia at Day 0 (LM), parasites/μl* | 49 | 5731.8 (4210.8, 7802.2) | 16 | 6762.3 (3811.7, 11997.0) | 0.621 | 8 | 12415 (5792, 14031) | 0.142 |
| PCT, hours | 48 | 47.5 (36.2, 59.8) | 18 | 54.2 (48.4, 62.0) | 0.035 | 8 | 60.3 (48.6, 73.0) | 0.019 |
| PC<sub>1/2</sub>, hours | 46 | 4.0 (3.4, 5.2) | 17 | 4.8 (4.0, 5.5) | 0.078 | 8 | 4.8 (4.2, 5.7) | 0.125 |
| Positive at Day 2: |  |  |  |  |  |  |  |  |
| LM | 49 | 16 (32.7) | 18 | 9 (50) | 0.193 | 8 | 5 (62.5) | 0.13 |
| qPCR | 46 | 19 (41.3) | 17 | 9 (52.3) | 0.409 | 8 | 5 (62.5) | 0.443 |
| Positive at Day 3: |  |  |  |  |  |  |  |  |
| LM | 49 | 2 (4.0) | 17 | 0 (0) | 1.000 | 8 | 0 (0) | 1.000 |
| qPCR | 48 | 7 (14.6) | 17 | 2 (11.8) | 1.000 | 8 | 1 (12.5) | 1.000 |
| **Ex vivo drug sensitivity (Day 0):** |  |  |  |  |  |  |  |  |
| CQ IC50, nM * | 13 | 41.5 (24.4-80.4) | 8 | 38.7 (10.4-108.0) | 0.856 | 5 | 51.3 (24.3-108.2) | 0.301 |
| **Candidate molecular markers of CQR (Day 0):** |  |  |  |  |  |  |  |  |
| *pvm**dr1* SNP: |  |  |  |  |  |  |  |  |
| Y976F | 47 | 34 (72.3) | 16 | 10 (62.5) | 0.459 | 6 | 3 (50.0) | 0.351 |
| F1076L | 47 | 45 (96) | 16 | 15 (93.8) | 1.000 | 6 | 6 (100) | 1.000 |
| *pvcrt* expression, R | 33 | 0.09 (0.05, 0.22) | 16 | 0.20 (0.15, 0.56) | 0.002 | 8 | 0.17 (0.14, 0.38) | 0.038 |
| *pvm**dr1* expression, R | 35 | 0.11 (0.04, 0.28) | 16 | 0.17 (0.06, 0.47) | 0.138 | 8 | 0.09 (0.05, 0.35) | 0.779 |

\*geometric mean (95% CI).
*p*-values correspond to results of Wilcoxon rank-sum test (for continuous variables) and Chi-square test (for proportions).
ACPR, adequate clinical and parasitological response; LM, light microscopy; CQR, chloroquine resistance; R, relative expression ratio.

### Drug-resistance characteristics

Phenotypic assessment of CQ susceptibility was conducted using the short-term schizont maturation assays at Day 0. A total of 41 cultures were conducted out of which 21 were valid for analysis (51% success rate). The overall CQ IC50 geometric mean was 40.4 nM (95% CI 31.5, 51.8; range 10.4-108.0). There were no differences in *ex vivo* CQ sensitivity between recurrent and non-recurrent infections (Table 4 and Fig 3).

**Fig 3.**
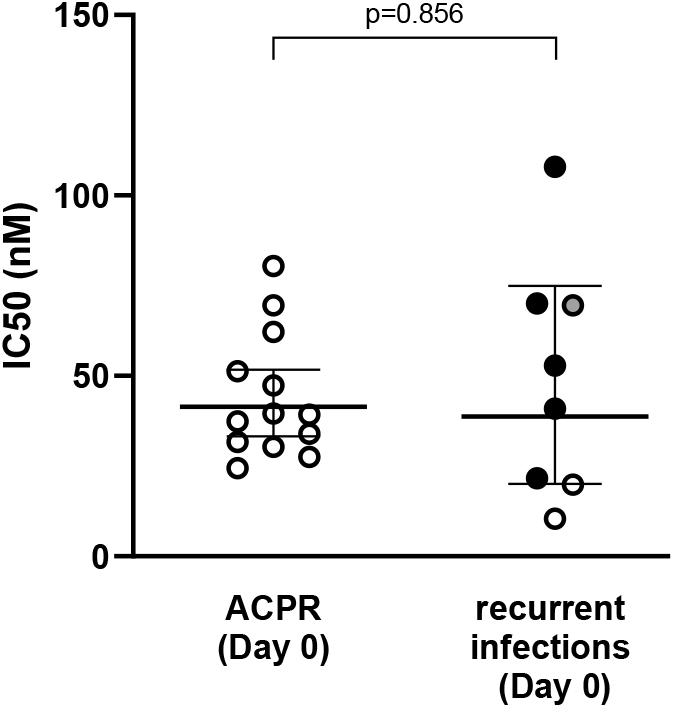
*Ex vivo* susceptibility to chloroquine in schizont maturation assays (SMA). Graph shows results of CQ-IC50 (nM) in Day 0 samples, stratified by treatment outcome. The type of recurrence based on identity-by-descent (IBD) analysis is indicated with colored dots (black, IBD-homologous recurrence; grey, IBD-heterologous recurrence; white, undetermined by WGS). *ACPR, adequate clinical and parasitological response*.

At genetic level, mutations in *pvmdr1* codons F976Y and F1076L were common (30.2% [19/63] for F976Y and 95.2% [60/63] for F1076L) but no differences were found in prevalence of mutations between treatment outcome groups at Day 0 (Table 4). In samples sequenced at DRec (n=10), F976Y and F1076L variants were found in 20% (2/10) and 100% (10/10) of the sequences, respectively.

Gene expression of *pvcrt* and *pvmdr1* was determined in all samples with sufficient RNA quantity at both Day 0 (n=51) and DRec (n=13). At Day 0, median *pvcrt* levels were 2.2-fold higher in infections that led to recurrence as compared to those with ACPR (p=0.002, Wilcoxon rank-sum), an increase that was also observed when only IBD-homologous recurrences were considered (Table 4). No difference between treatment outcome groups at Day 0 was found for *pvmdr1* levels (p=0.138; Table 4). Comparisons at Day 0 were not affected after adjustment by multiple comparisons (Fig 4). We did not find an association between ring-stage proportion and gene expression of either *pvcrt* in linear regression models to suggest there was a potential confounding effect of parasite life-stage on transcript levels (see Fig S3). There was no correlation between parasite *ex vivo* CQ susceptibility (defined by IC50 values) and gene expression of *pvcrt* (n=13; Spearman’s rho=-0.217, p=0.476) or *pvmdr1* (n=17; Spearman’s rho=-0.120, p=0.646).

**Fig 4.**
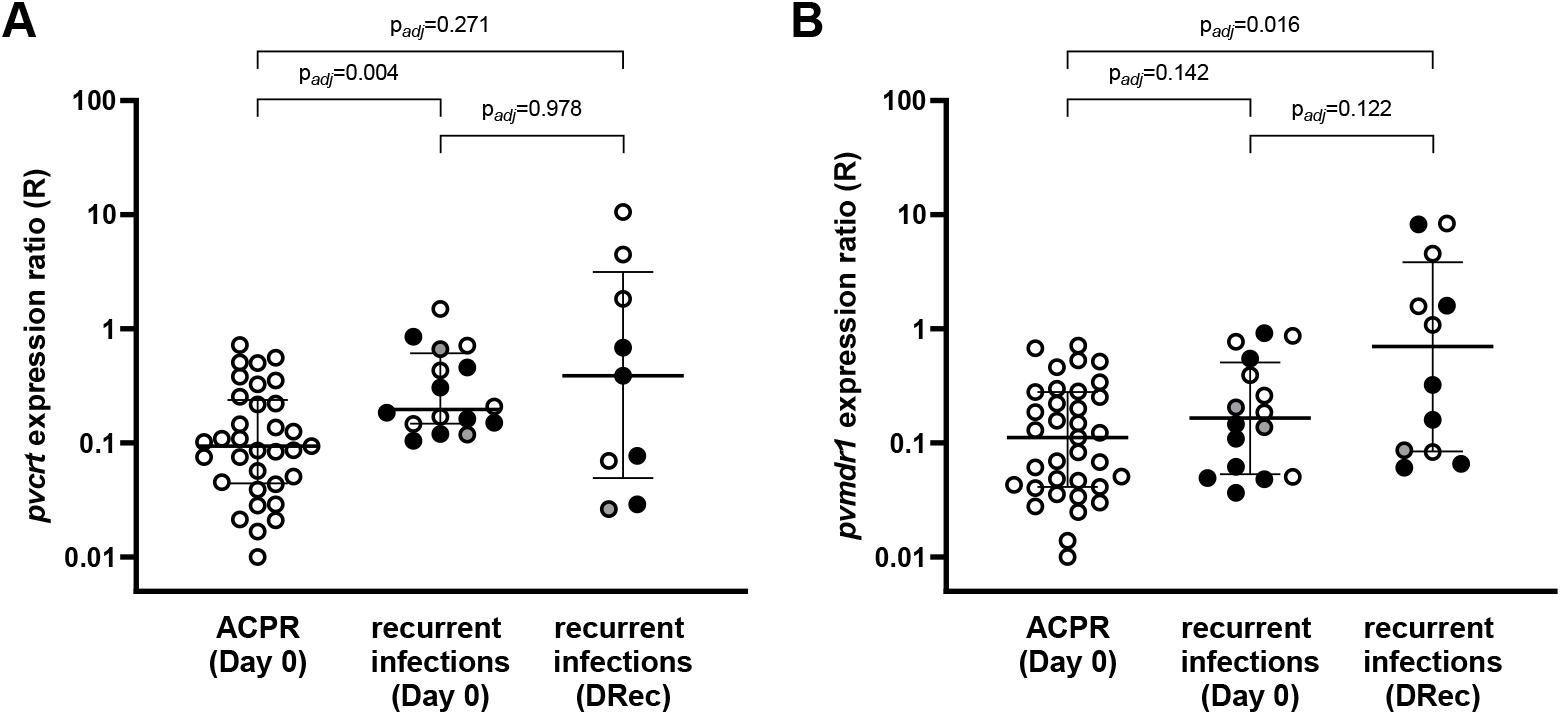
Gene expression of *pvcrt* and *pvmdr1*. Expression ratio R of *pvcrt* (A) or *pvmdr1* (B) relative to reference gene *β-tubulin* is displayed in log-scale. Samples are stratified by treatment outcome and day of collection. Horizontal lines indicate median and IQR. The type of recurrence based on identity-by-descent (IBD)analysis is indicated with colored dots (black, IBD-homologous recurrence; grey, IBD-heterologous recurrence; white, undetermined by WGS). Adjusted p-values from pairwise Wilcoxon rank-sum test with Benjamini-Hochberg correction for multiple testing are reported. *ACPR, adequate clinical and parasitological response*.

Parasites collected at DRec showed a wide range of *pvcrt* expression levels (n=9; R=0.39 [IQR 0.07, 1.83]), which did not differ significantly from those in the ACPR group (p_adj_=0.271, rank-sum; Fig 4) or in Day 0 samples with a later recurrence, either in unpaired (p_adj_=0.978) or in Day0/DRec matched pairs analysis (n=8; ΔR[DRec-Day0]=-0.1 [IQR −0.6, 0.8], p=1.000, signed-rank test; see Fig S4). Expression of *pvmdr1* was higher at DRec (n=12; R=0.70 [IQR 0.08, 3.09]) as compared to ACPR (p_adj_=0.016; Fig 4), but not compared to Day 0 samples with a later recurrence, either in unpaired (p_adj_=122) or Day0/DRec matched pairs analysis (n=10; ΔR=0.2 [IQR −0.1, 1.3], p=0.203; Fig S4).

## DISCUSSION

This study evaluated CQ therapeutic efficacy for uncomplicated *P. vivax* malaria in Gia Lai province, Vietnam. The success rate for *P. vivax* treatment was 100% by Day 28 and 75.4% by Day 42. All recurrences detected by qPCR (n=18) occurred after day 33 in the presence CQ levels below MEC (<100 mg/ml) and were associated with longer PCT of primary infection, younger age, and higher transcript levels of *pvcrt* at baseline. Eighty percent of recurrences (8/10) with WGS data were homologous to primary infections using IBD analysis, leading to a homologous recurrence rate at Day 42 of at least 44% (8/18).

Among previous CQ TES studies conducted in Vietnam that detected early recurrences (*i*.*e*. <Day 28) (20,21,23), only one confirmed recurrent infections (n=3) with blood CQ concentration above MEC in Quang Nam in 2011 (20). Although the present study was conceived as a continuation of CQR monitoring in Quang Nam province (20), malaria cases rapidly decreased there after 2013, with mostly residual *P. vivax* by 2015, which was insufficient to conduct classical TES designs due to obvious time and logistic constraints (48). The primary endpoints presented in this study suggest CQ remained largely efficacious in Gia Lai province until 2017, with mostly late recurrences detected after Day 28 in the presence of blood CQ concentrations below MEC, similar to findings in the southern province of Binh Phuoc (22).

Recurrences occurring after Day 28 of follow-up and in the presence of blood CQ concentrations below MEC have generally been classified as relapses (4,49), which is in line with the frequent relapse phenotype of *P. vivax* strains in the study region (*i*.*e*. mean time-to-relapse of 41 days (3)). Nevertheless, a thorough revision of *P. vivax* relapses in tropical areas also showed that recrudescent infections of low-grade CQR parasites will become patent at the same time as relapsing infections, due to parasite expansion coincident with the fall in drug levels (7). Some of the findings in the present study undoubtedly suggest that low-grade *P. vivax* CQR should not be ruled out.

First, recurrences had a significantly longer PCT than ACPR infections. The difference in PCT was independent of parasite densities at Day 0, suggesting that they were not attributable to an incomplete clearance of a higher parasite inoculum (7). The observation that recurrences occurred in younger individuals (*i*.*e*. less frequently exposed to *P. vivax* infection throughout life [7, 51], hence with weaker immune response to support drug-mediated clearance) could also have further contributed to the persistence of low-grade CQR parasites. Regardless, given that the median age in both treatment outcome groups falls within early adulthood (18 vs 25 years old) the effect of age, if any, would probably be small.

Second, genetic analysis revealed a remarkably high proportion of homologous recurrences using both common microsatellite genotyping (80% rate using 4 MS marker) but also fine-scale genomic approaches, which resulted in a minimum of 44% (8/18) homologous recurrence rate with a degree of relatedness to Day 0 infections above 98% IBD. An earlier review of *P. vivax* clinical studies concluded that relapses in endemic areas commonly occur with heterologous genotypes (as over-repeated liver inoculations lead to the accumulation of genetically diverse hypnozoites), whereas homologous relapses are more likely to occur either in patients recently treated with PQ (*i*.*e*. those that cleared pre-existing hypnozoites) or in those living in areas with very little parasite genetic diversity (7). More recently, a study conducted in Cambodia investigated recurrent *P. vivax* infections among 20 patients reallocated to an area with no malaria transmission, in order to exclude the possibility of acquiring new infections(10). Although recurrences were comparable to our study in terms of recurrence time (>Day 33) and blood CQ concentrations (<MEC), they were classified as genetically heterologous based on allelic differences by WGS or high-throughput SNP genotyping (6/8), in contrast to the low heterologous rate in our study despite using an IBD approach (2/10). The authors concluded that all recurrences were thus caused by relapses (10). The possibility that IBD-homologous recurrences we identified in Vietnam were also caused by relapses (or even reinfections) with an identical parasite genotype cannot be totally excluded, as we could be underestimating the presence of new alleles due to a lower WGS coverage in our study. However, the latter seems unlikely given: i) the excess of genetically identical parasites in recurrent samples observed using IBD (low genome coverage would underestimate relatedness by IBD compared to IBS analysis, which can misclassify unrelated parasites as homologous due to sharing of alleles by chance); ii) the fact that the number of genetically different sample pairs remained when only the dominant clones were considered in the Cambodian study (thus increased depth does not explain the differences in heterologous rate between studies); iii) the higher rate of polyclonally among primary infections with a later recurrence (89% [16/18]; mean COI=3 by MS) as compared to 50% (10/20) in Cambodia (suggesting a higher genetic diversity in Vietnam that would increase the chance of detecting heterologous recurrences if those were caused by a relapse; indeed, the very limited use of PQ as radical cure in Vietnam may further favor high hypnozoite diversity in the country (50)).

Finally, the third finding suggesting some degree of CQR is the higher baseline transcript levels of *pvcrt* in recurrent infections compared to patients with ACPR, independently of parasite stage composition (51,52). This suggests that *P. vivax* parasites expressing higher levels of *pvcrt* may have reduced CQ susceptibility. Data on *pvcrt* expression in response to CQ treatment is limited. In line with our results, a previous study in Brazil found increased *pvcrt* (and *pvmdr1*) levels in *P. vivax* parasites with CQR phenotype both at Day0 and at time of recurrence (17). A recent study in non-human primates infected with a genetic cross of *P. vivax* parasites with different CQ responses identified a strong selection of the chromosome 1 region that includes *pvcrt*, linked to the CQR phenotype and upregulation of *pvcrt* expression(18), supporting a role of *pvcrt* in the molecular mechanism of CQR. As for *pvmdr1*, increased expression found at DRec (although not confirmed in the matched pairs analysis) could hypothetically reflect a mechanism of drug resistance or be a consequence of an altered membrane transport after drug exposure, but current knowledge of the physiological function of *pvmdr1* is still very limited (53). There is a clear need to further investigate the patterns of *pvcrt* and *pvmdr1* expression in a higher number of ACPR and recurrent samples (both homologous and heterologous) to elucidate their role in CQ susceptibility.

Considering a genetically homologous recurrence as a true recrudescence would imply that parasites observed at Day 0 remain in the human host at levels below qPCR detection limit for several weeks while CQ blood concentrations are over MEC. It has been proposed that *P. vivax* parasites seen in the bloodstream represent only a portion of the total parasite biomass, and that a fraction of parasites may hide in the bone marrow (54–56). Several studies strongly support the role of this tissue as a parasite reservoir, both in human and non-human primates (57)(58). An hypothetical scenario could be one in which a sub-populations of parasites with higher *pvcrt* expression persist in bone marrow parenchyma where CQ levels are lower, leading to homologous recurrences as soon as drug concentrations decrease significantly below the MEC threshold, furthermore coinciding with the time hypnozoite stages are reactivated producing relapse (7).

A limitation of this study was the impossibility to characterize the phenotype of isolates at time of recurrence to provide further evidence of low-grade CQR recrudescence in addition to our findings at the genomic level. Additionally, for the number of samples eligible for *ex vivo* assays or genomic analysis were lower than the total number of patients recruited, a common challenge of *P. vivax* field studies (*i*.*e*. low parasite density infections and poor adaptation to *ex vivo* conditions). Another difficulty is the intrinsic logistical constraints linked to TES in settings where transmission is being drastically reduced, which questions the feasibility of classical *in vivo* studies under these epidemiological conditions. Such limitation could potentially be overcome once a CQR marker is validated and can be implemented into molecular surveillance, similarly to the ongoing surveillance of *P. falciparum* artemisinin resistance in Greater Mekong Subregion based on the *pfkelch-13* gene (59).

In conclusion, despite the full efficacy of *P. vivax* CQ treatment at Day 28, we find clinical (PCT), genomic (high rate of IBD homology) and transcriptional (increased *pvcrt* expression) evidence suggesting recurrences after Day 28 can be attributable to low-grade CQR recrudescence rather than genetically homologous relapses of liver hypnozoites. Emergence of CQR parasites should be closely monitored. The conclusive identification of a molecular marker is of uttermost importance to unequivocally define *P*.*vivax* CQR, and detect early warning signs of CQR in molecular surveillance studies conducted in endemic areas. The finding that *pvcrt* (and even *pvmdr1*) may play a role in *in vivo* CQ susceptibility deserves further attention.

## Data Availability

The datasets generated and/or analysed during the current study are included in this published article and/or are available from the corresponding author on reasonable request.

## ACKNOWLEDGEMENTS

We sincerely thank all individuals that accepted to enroll in the trial, all staff from Chu R’Cam CHC and local authorities in Krong Pa district. We are especially grateful to Tran Tuyet Mai, Nguyen Thi Thuy Duong and Do Manh Ha for their support to field work; and to Ric Price and Kamala Thriemer (Menzies School of Health Research) for clinical trial protocol guidelines and helpful discussion.

The study was funded by Belgium Development Cooperation (DGD) under the Framework Agreement Program between DGD and ITM (FA3-III, 2014-2016, FA4 2017-2021). This publication uses data from the MalariaGEN SpotMalaria Project, coordinated by the MalariaGEN Resource Centre supported by Wellcome (098051, 090770). Both Chloroquine and Primaquine were provided at no cost by Sanofi.

## AUTHORS CONTRIBUTIONS

Conceived and designed the study: ERV, NVH, AE, ARU; performed the experiments: ERV, NVH, JHK, NTS, NTTH, NTHN, PG, ES; analyzed the data: ERV, NVH, JHK; contributed data collection/logistics/materials/analysis tools: NLH, TTM, NTTD, NDL, TTD, NXX, AE; wrote the paper: ERV, JHK, ARU. All authors read and approved the final version of the manuscript. The authors declare they have no conflicts of interest.

## Supplemental Material

**Table S1.**
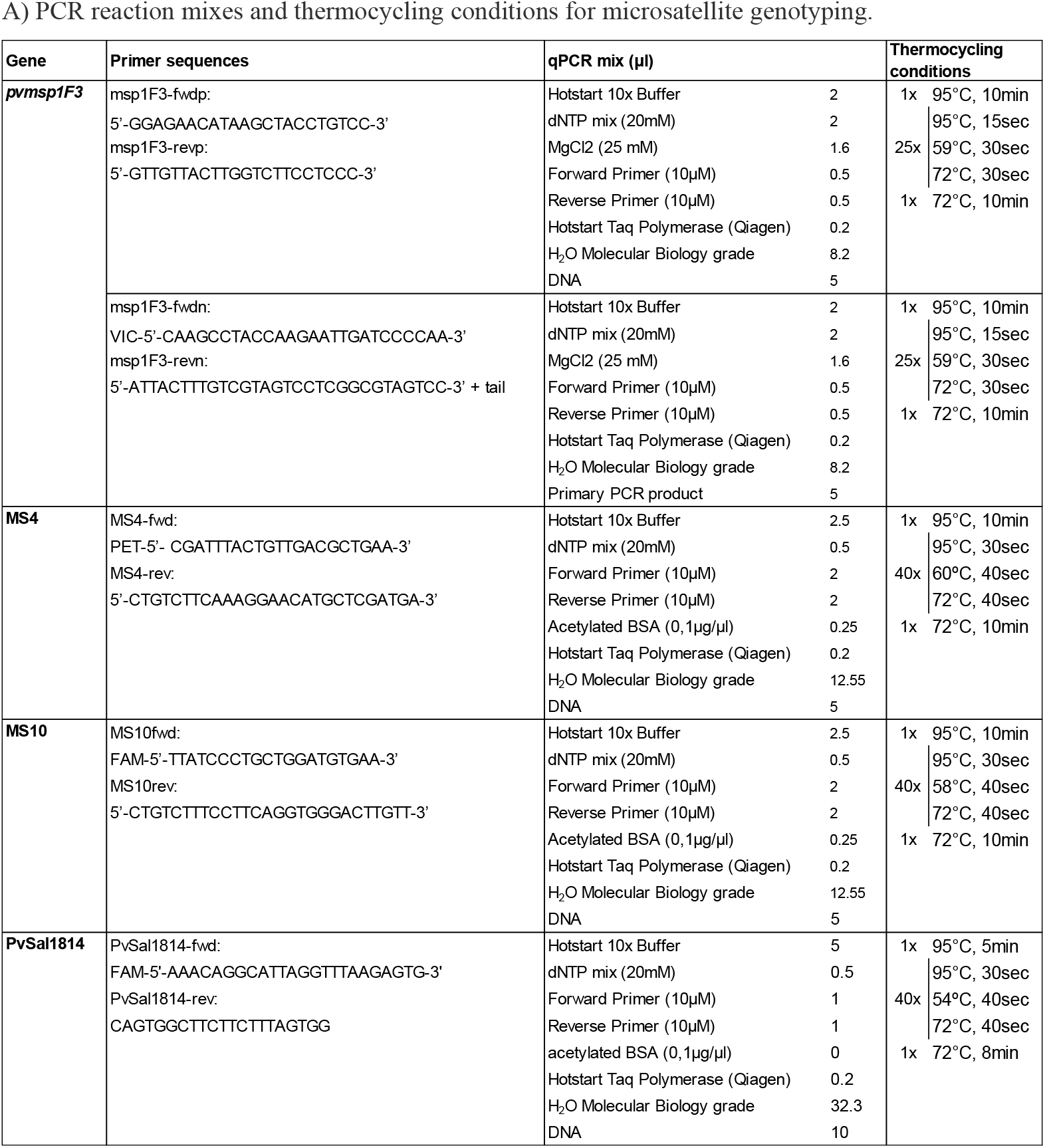
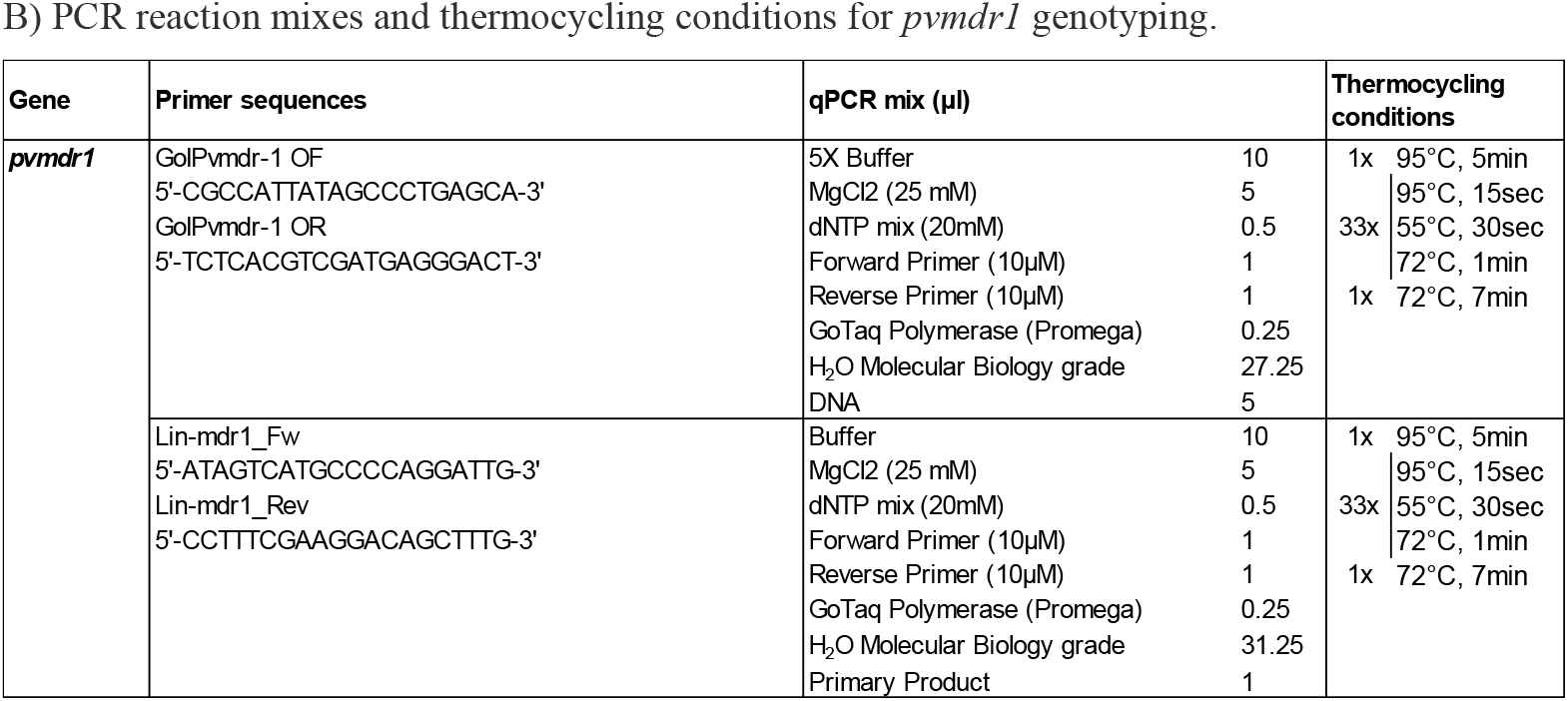
PCR reaction mixes and thermocycling conditions.

**Data S1** (Excel [S1_Data_GRC.xlsx]). Genetic Report Card (SpotMalaria). The dataset contains drug-resistance genotypes as well as the SNP barcode (Sheet A). An explanatory table with detail of sequenced positions and reference/mutant codon is provided in Sheet B.

**Table S2.** Whole-genome sequencing coverage.

| Patient code | Day 0 |  |  | Day of recurrence |  |  |
| --- | --- | --- | --- | --- | --- | --- |
| | parasites/μl | coverage (mean X ± SD) | % missing positions | parasites/μl | coverage (mean X ± SD) | % missing positions |
| 007 | 41506 | 41.2 ± 52 | 0.8% | 769 | 21.4 ± 33 | 4.4% |
| 008 | 6068 | 27.6 ± 51 | 13.8% | 3428 | 45.0 ± 43 | 0.4% |
| 010 | 5431 | 6.8 ± 11 | 0.9% | 212 | 1.6 ± 15 | 88.7% |
| 018 | 2408 | 25.5 ± 37 | 2.9% | 103 | 3.3 ± 22 | 81.9% |
| 022 | 13430 | 38.9 ± 54 | 1.6% | 444 | 44.0 ± 46 | 0.4% |
| 027 | 5517 | 37.1 ± 57 | 4.6% | 3268 | 42.2 ± 42 | 0.4% |
| 034 | 4010 | 42.1 ± 56 | 2.1% | 1536 | 45.5 ± 47 | 0.4% |
| 037 | 11401 | 38.5 ± 55 | 2.5% | 310 | 37.5 ± 41 | 0.6% |
| 043 | 55892 | 42.7 ± 39 | 0.3% | 7477 | 34.2 ± 40 | 0.8% |
| 044 | 274 | 46.4 ± 51 | 0.6% | 68 | 2.5 ± 9 | 48.6% |
| 047 | 4927 | 25.0 ± 55 | 27.8% | 91 | 35.0 ± 39 | 0.6% |
| 048 | 13965 | 50.1 ± 47 | 0.3% | 856 | 16.9 ± 41 | 35.5% |
| 053 | 11262 | 8.6 ± 40 | 81.0% | 2469 | 21.8 ± 52 | 34.4% |
| 054 | 14096 | 32.3 ± 50 | 4.0% | 440 | 35.7 ± 55 | 3.9% |
| **All** |  | **33.1 ± 47** | **10.2%** |  | **27.6 ± 38** | **21.5%** |

**Fig S1.**
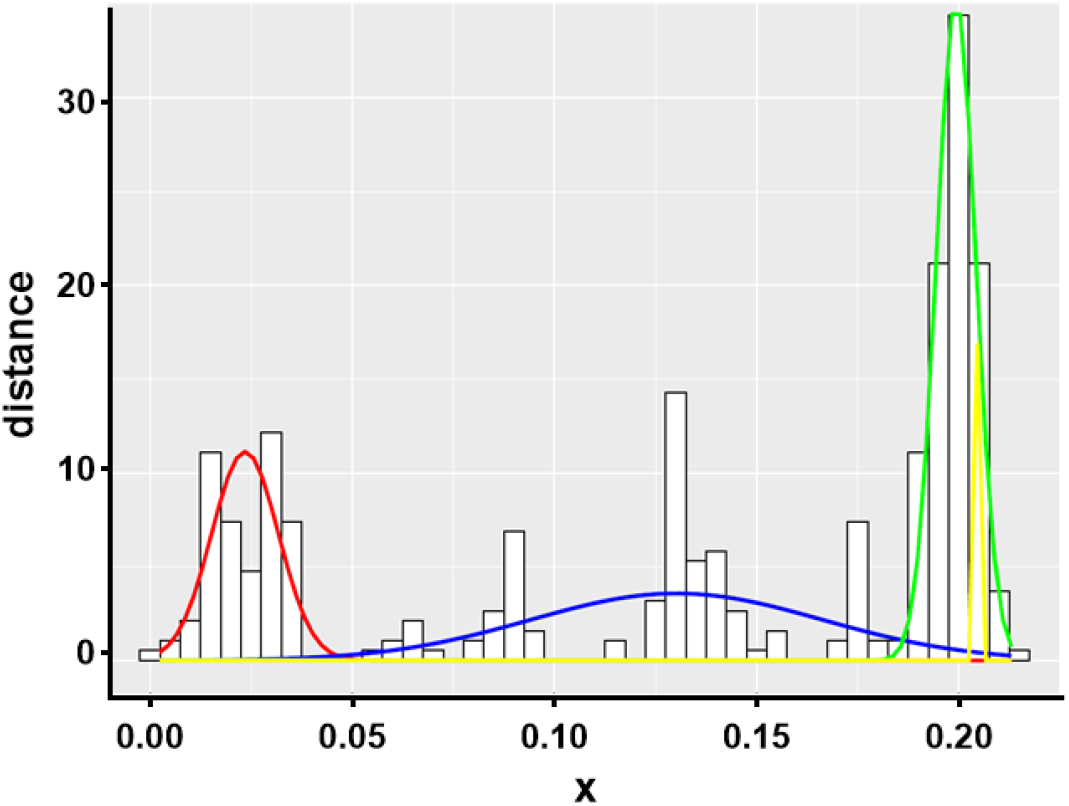
Distribution of discordant SNPs by whole-genome sequencing. The proportion of discordant SNPs was determined by calculating the Prevosti distance between all pair-wise comparisons (*i*.*e*. number of allelic differences/number of possible differences) using the R-package *poppr* (https://cran.r-project.org/web/packages/poppr/index.html). Due to the high likelihood of multiclonal infections in *P. vivax*, distances were fitted as the sum of four Gaussian distributions using the R-package *mixtools* (https://cran.r-project.org/web/packages/mixtools/index.html). The distribution with the lowest proportion of discordant SNPs (red line) was used to define the threshold (mean plus 3x the standard deviation for that distribution) of homologous recurrence for identify-by-state (IBS) analysis.

**Fig S2.**
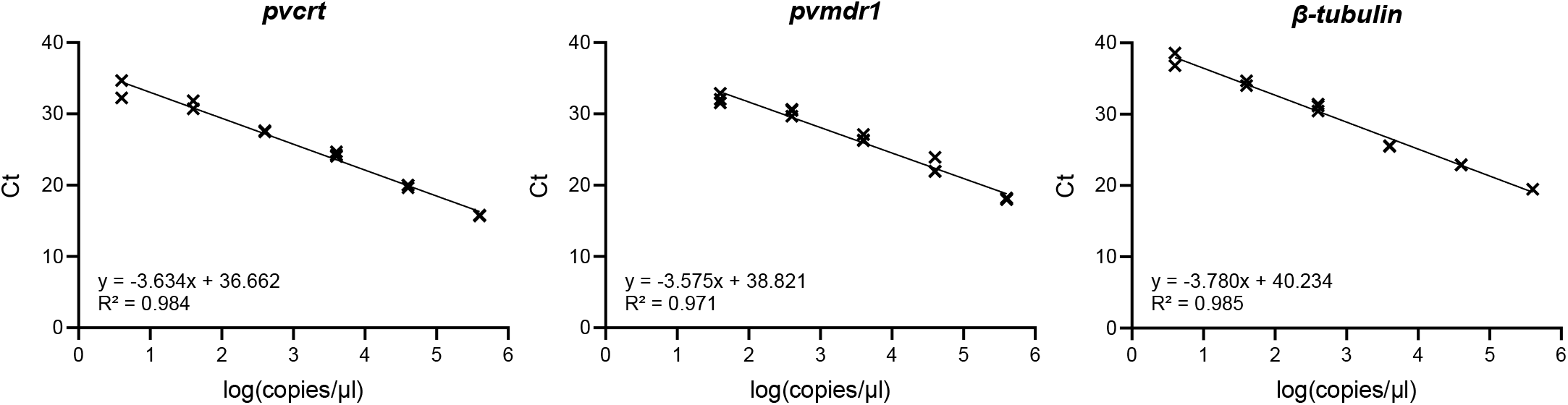
qPCR standard curves for *pvcrt, pvmdr1* and *β-tubulin*. Standard curves were prepared using 10-fold serial dilutions of dsDNA fragments covering the targeted region in each gene of interest (range 4000000-4 copies/µl). Amplification efficiencies calculated from cycle threshold (Ct) *versus* dsDNA copy number (log transformed) plots were 1.88, 1.90 and 1.84 for *pvcrt, pvmdr1* and *β-tubulin*, respectively.

**Table S3.** Microsatellites genotyping. Results of allele calling using Genetools software. Shared alleles between sample pairs at Day 0 and Day of recurrence (DRec) are indicated in bold.

| Patient<br>code | Day | PvMSP1.F3 |  |  |  |  | MS4 |  | MS10 |  | PvSal1814 |  |  |  |  |  |
| --- | --- | --- | --- | --- | --- | --- | --- | --- | --- | --- | --- | --- | --- | --- | --- | --- |
|  |  | Allele1 | Allele2 | Allele3 | Allele4 | Allele5 | Allele1 | Allele2 | Allele1 | Allele2 | Allele1 | Allele2 | Allele3 | Allele4 | Allele5 | Allele6 |
| 003 | Day0 | 264 | 313 |  |  |  | 195 | 198 | 216 |  | 528 | 543 | 558 |  |  |  |
|  | DRec | 264 |  |  |  |  |  | 198 |  |  | 528 | 543 |  |  |  |  |
| 007 | Day0 | 229 |  | 264 | 278 | 313 | 198 | 201 | 201 | 207 | 599 | 614 | 629 |  |  |  |
|  | DRec |  | 259 |  |  | 313 |  | 201 | 201 |  | 599 | 614 | 629 |  |  |  |
| 008 | Day0 | 276 |  |  |  |  | 195 |  | 201 |  | 563 | 578 |  |  |  |  |
|  | DRec | 276 | 325 |  |  |  | 195 |  | 201 |  | 563 | 578 |  |  |  |  |
| 010 | Day0 | 259 | 306 |  |  |  | 204 |  | 213 |  |  |  |  |  |  |  |
|  | DRec |  |  |  |  |  |  |  |  |  |  |  |  |  |  |  |
| 018 | Day0 | 259 | 264 | 316 |  |  | 198 | 201 | 204 | 213 | 510 | 525 | 540 |  |  |  |
|  | DRec | 259 |  |  |  |  |  |  | 204 |  |  |  |  |  |  |  |
| 021 | Day0 | 253 |  | 300 |  |  |  | 204 | 195 |  |  |  |  | 573 | 588 | 603 |
|  | DRec |  | 259 |  |  |  | 201 |  |  | 207 | 528 | 543 | 558 |  |  |  |
| 022 | Day0 | 219 | 278 | 327 |  |  | 198 |  | 195 |  | 599 | 614 | 629 |  |  |  |
|  | DRec |  | 278 |  |  |  | 198 |  | 195 |  | 599 |  |  |  |  |  |
| 027 | Day0 |  |  | 325 | 374 |  | 201 |  | 198 |  | 555 | 584 | 599 |  |  |  |
|  | DRec | 147 | 197 |  | 374 |  | 201 |  | 198 |  | 555 | 584 | 599 |  |  |  |
| 028 | Day0 | 147 | 197 | 325 | 374 |  | 201 |  | 198 |  |  |  |  |  |  |  |
|  | DRec |  |  |  |  |  |  |  |  |  |  |  |  |  |  |  |
| 034 | Day0 | 325 |  |  |  |  | 201 |  | 198 |  |  | 581 |  | 596 |  |  |
|  | DRec | 325 |  |  |  |  | 201 |  | 198 |  | 555 |  | 584 |  |  |  |
| 037 | Day0 | 259 |  |  |  |  | 201 |  | 201 |  |  |  |  |  |  |  |
|  | DRec |  |  |  |  |  |  |  |  |  |  |  |  |  |  |  |
| 043 | Day0 | 264 | 272 |  |  |  |  |  | 204 |  | 555 | 569 | 584 |  |  |  |
|  | DRec |  | 272 |  |  |  | 198 |  | 204 |  |  |  | 584 | 614 |  |  |
| 044 | Day0 | 264 |  |  |  |  | 204 |  | 210 |  | 584 | 599 | 614 |  |  |  |
|  | DRec | 264 |  |  |  |  |  |  | 210 |  | 584 | 599 | 614 |  |  |  |
| 047 | Day0 | 253 | 264 |  |  |  | 195 | 201 | 207 |  | 599 |  |  |  |  |  |
|  | DRec | 253 | 264 |  |  |  |  | 201 | 204 |  |  |  |  |  |  |  |
| 048 | Day0 | 264 |  |  |  |  | 223 |  | 201 |  |  |  |  |  |  |  |
|  | DRec | 264 |  |  |  |  | 223 |  | 201 |  | 599 |  |  |  |  |  |
| 050 | Day0 | 264 |  |  |  |  | 198 |  | 213 |  | 620 | 635 |  |  |  |  |
|  | DRec | 264 |  |  |  |  |  |  | 213 |  | 620 |  |  |  |  |  |
| 053 | Day0 | 258 |  |  |  |  | 201 |  | 201 |  |  | 573 | 588 |  |  |  |
|  | DRec | 258 |  |  |  |  | 201 |  | 201 |  | 558 | 573 | 588 | 603 |  |  |
| 054 | Day0 | 250 |  |  |  |  | 213 |  | 204 |  | 620 | 635 |  |  |  |  |
|  | DRec | 250 |  |  |  |  | 213 |  | 204 |  | 620 | 635 |  |  |  |  |

**Table S4.**
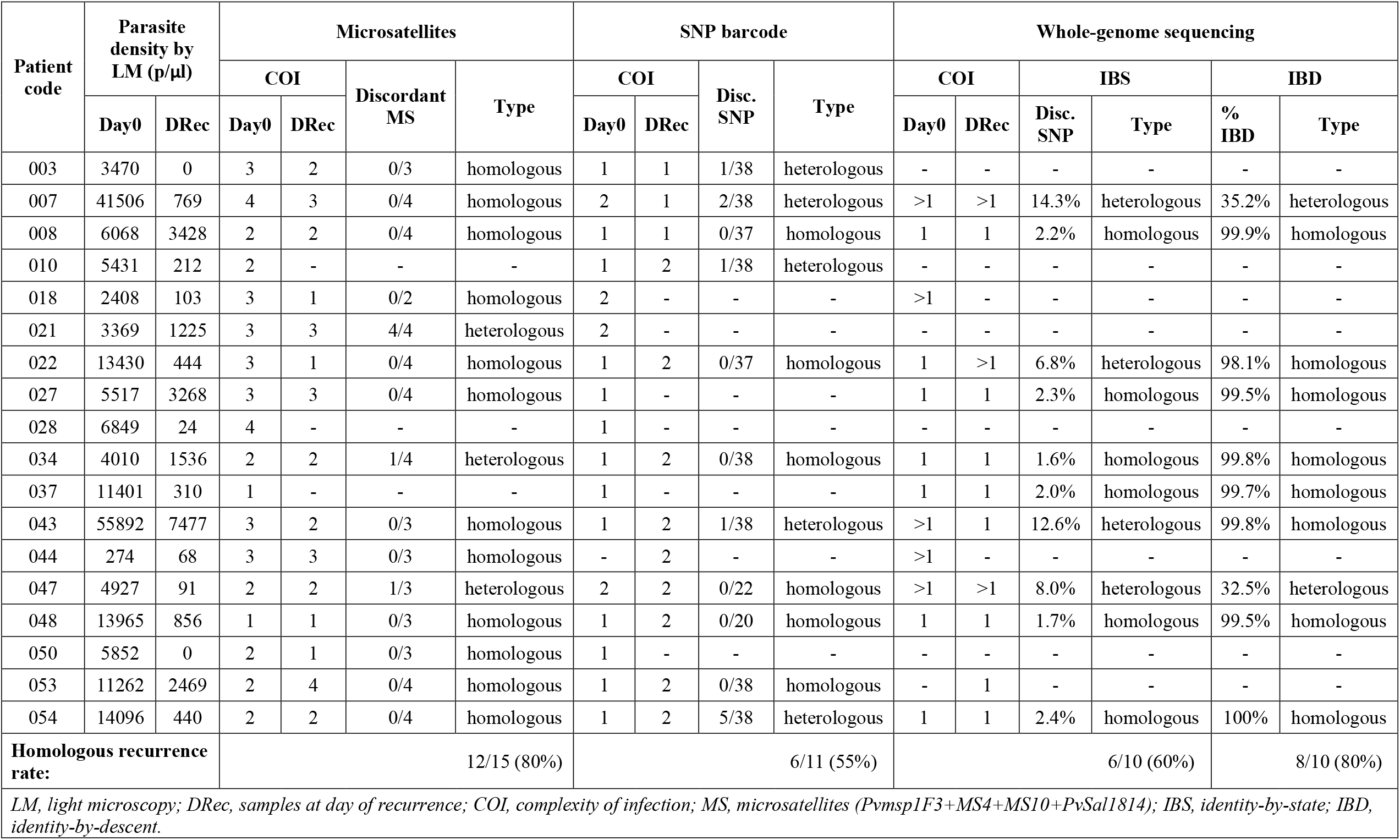
Genotyping and categorization of recurrent infections.

| Patient<br>code | Parasite<br>density by<br>LM (p/μl) |  | Microsatellites |  |  |  | SNP barcode |  |  |  | Whole-genome sequencing |  |  |  |  |  |
| --- | --- | --- | --- | --- | --- | --- | --- | --- | --- | --- | --- | --- | --- | --- | --- | --- |
|  |  |  | COI |  | Discordant<br>MS | Type | COI |  | Disc.<br>SNP | Type | COI |  | IBS |  | IBD |  |
|  | Day0 | DRec | Day0 | DRec |  |  | Day0 | DRec |  |  | Day0 | DRec | Disc.<br>SNP | Type | %<br>IBD | Type |
| 003 | 3470 | 0 | 3 | 2 | 0/3 | homologous | 1 | 1 | 1/38 | heterologous | - | - | - | - | - | - |
| 007 | 41506 | 769 | 4 | 3 | 0/4 | homologous | 2 | 1 | 2/38 | heterologous | >1 | >1 | 14.3% | heterologous | 35.2% | heterologous |
| 008 | 6068 | 3428 | 2 | 2 | 0/4 | homologous | 1 | 1 | 0/37 | homologous | 1 | 1 | 2.2% | homologous | 99.9% | homologous |
| 010 | 5431 | 212 | 2 | - | - | - | 1 | 2 | 1/38 | heterologous | - | - | - | - | - | - |
| 018 | 2408 | 103 | 3 | 1 | 0/2 | homologous | 2 | - | - | - | >1 | - | - | - | - | - |
| 021 | 3369 | 1225 | 3 | 3 | 4/4 | heterologous | 2 | - | - | - | - | - | - | - | - | - |
| 022 | 13430 | 444 | 3 | 1 | 0/4 | homologous | 1 | 2 | 0/37 | homologous | 1 | >1 | 6.8% | heterologous | 98.1% | homologous |
| 027 | 5517 | 3268 | 3 | 3 | 0/4 | homologous | 1 | - | - | - | 1 | 1 | 2.3% | homologous | 99.5% | homologous |
| 028 | 6849 | 24 | 4 | - | - | - | 1 | - | - | - | - | - | - | - | - | - |
| 034 | 4010 | 1536 | 2 | 2 | 1/4 | heterologous | 1 | 2 | 0/38 | homologous | 1 | 1 | 1.6% | homologous | 99.8% | homologous |
| 037 | 11401 | 310 | 1 | - | - | - | 1 | - | - | - | 1 | 1 | 2.0% | homologous | 99.7% | homologous |
| 043 | 55892 | 7477 | 3 | 2 | 0/3 | homologous | 1 | 2 | 1/38 | heterologous | >1 | 1 | 12.6% | heterologous | 99.8% | homologous |
| 044 | 274 | 68 | 3 | 3 | 0/3 | homologous | - | 2 | - | - | >1 | - | - | - | - | - |
| 047 | 4927 | 91 | 2 | 2 | 1/3 | heterologous | 2 | 2 | 0/22 | homologous | >1 | >1 | 8.0% | heterologous | 32.5% | heterologous |
| 048 | 13965 | 856 | 1 | 1 | 0/3 | homologous | 1 | 2 | 0/20 | homologous | 1 | 1 | 1.7% | homologous | 99.5% | homologous |
| 050 | 5852 | 0 | 2 | 1 | 0/3 | homologous | 1 | - | - | - | - | - | - | - | - | - |
| 053 | 11262 | 2469 | 2 | 4 | 0/4 | homologous | 1 | 2 | 0/38 | homologous | - | 1 | - | - | - | - |
| 054 | 14096 | 440 | 2 | 2 | 0/4 | homologous | 1 | 2 | 5/38 | heterologous | 1 | 1 | 2.4% | homologous | 100% | homologous |
| Homologous recurrence rate: |  |  | 12/15 (80%) |  |  |  | 6/11 (55%) |  |  |  | 6/10 (60%) |  |  |  | 8/10 (80%) |  |
| LM, light microscopy; DRec, samples at day of recurrence; COI, complexity of infection; MS, microsatellites (Pvmsp1F3+MS4+MS10+PvSal1814); IBS, identity-by-state; IBD, identity-by-descent. |  |  |  |  |  |  |  |  |  |  |  |  |  |  |  |  |

**Fig S3.**
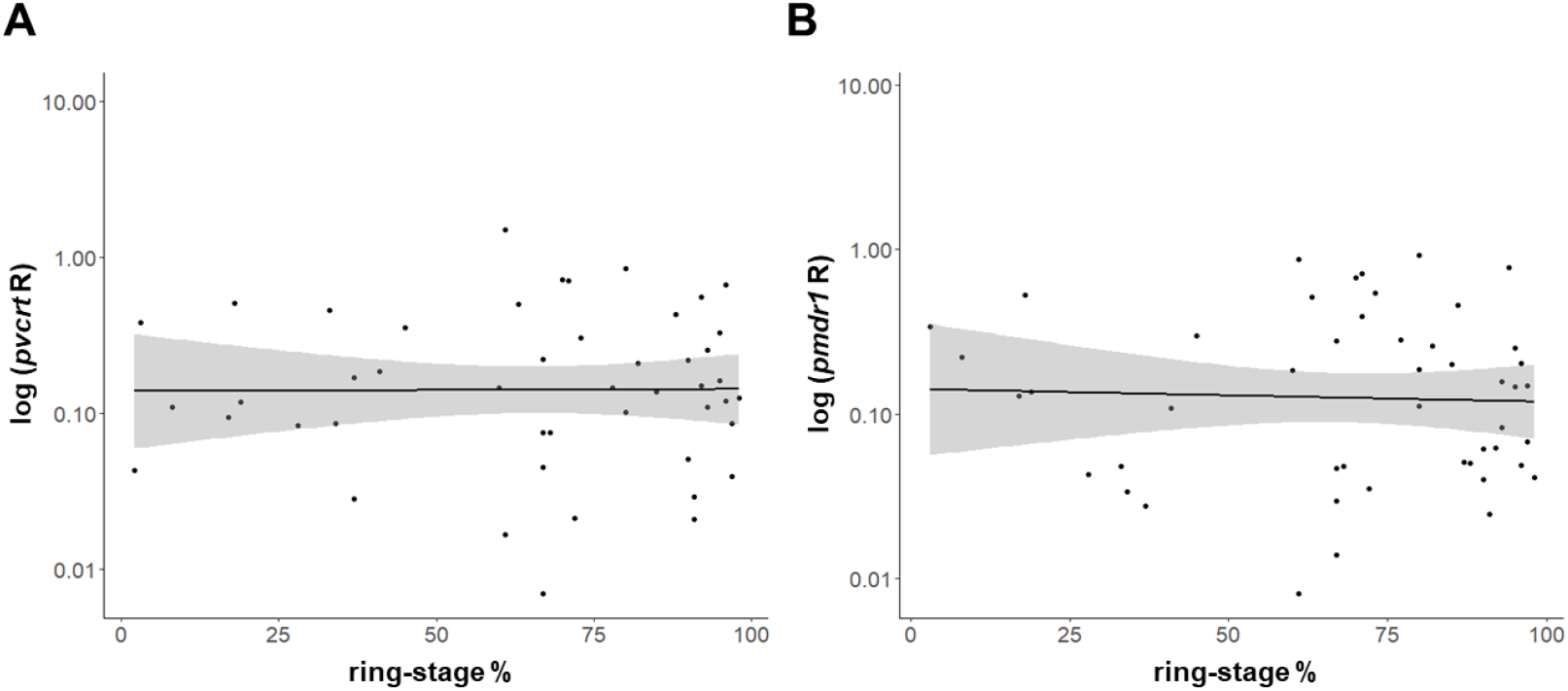
Gene expression of *pvcrt* and *pvmdr1* and proportion of ring-stages in infections at Day 0. Gene expression ratios (R) of *pvcrt* (A) and *pvmdr1* (B) relative to reference gene *β-tubulin* are displayed in log-scale (y axis). Linear regression was fitted to data (black line with standard error bounds in grey) did not show an association between R and ring-stage percentage (*pvcrt*: coef.=-0.0003, p=0.886; *pvmdr1*: coef.=-0.001, p=0.655).

**Fig S4.**
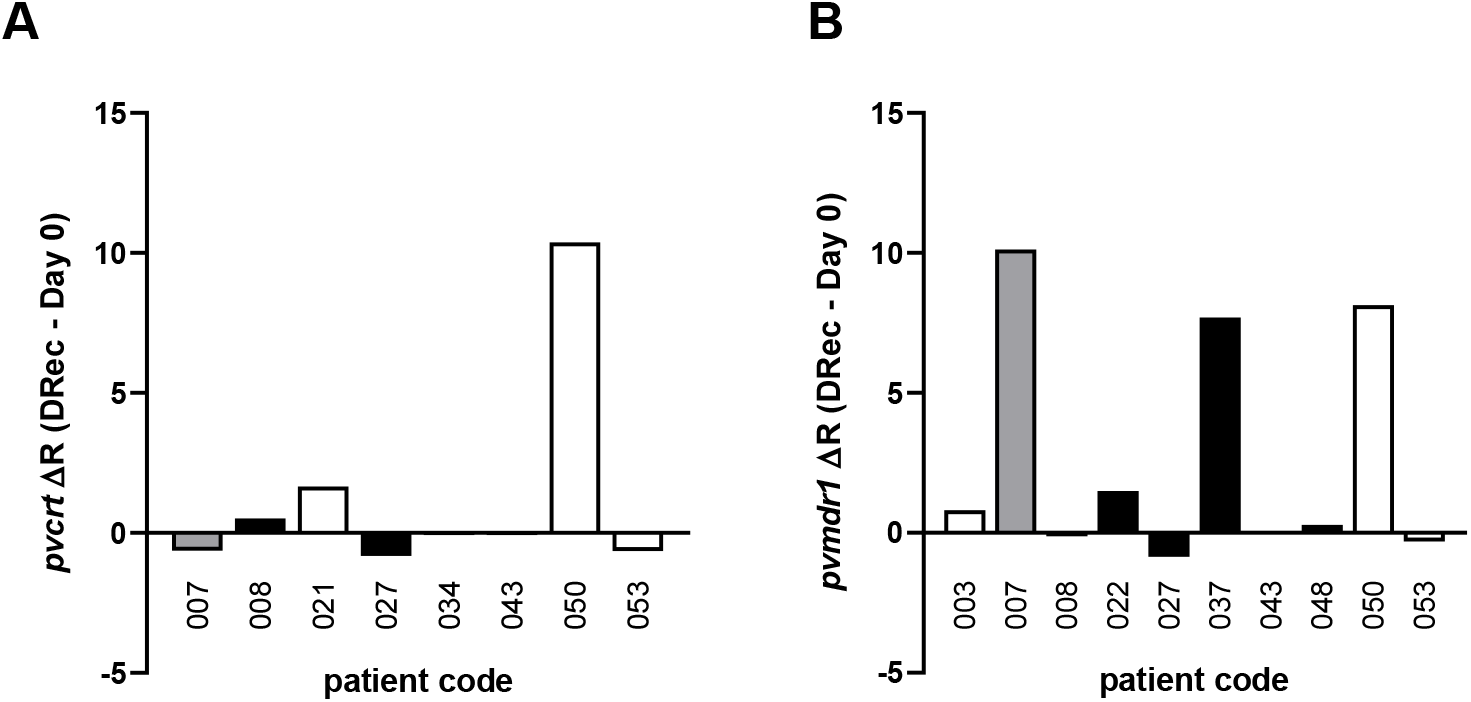
Gene expression of *pvcrt* and *pvmdr1* in Day 0/DRec paired samples. Differences in gene expression ratio R between infections at DRec and paired infections at Day 0 are shown for both *pvcrt* (A) and *pvmdr1* (B). Type of recurrences based on identity-by-descent (IBD) analysis is shown with coloured bars (black, IBD-homologous recurrence; grey, IBD-heterologous recurrence; white, undetermined by WGS).

## Notes

### Competing Interest Statement

The authors have declared no competing interest.

### Clinical Trial

NCT02610686

### Author Declarations

The study received approval from ethics committee of the NIMPE-Ministry of Health, Hanoi, Vietnam (351/QD-VSR and QD2211/QD-BYT); the Institutional Review Board of the Institute of Tropical Medicine, Antwerp, Belgium (937/14); and the ethics committee of Antwerp University Hospital (UZA), Antwerp, Belgium (14/15/183).

